# Benchmarking Mendelian Randomization methods for causal inference using genome-wide association study summary statistics

**DOI:** 10.1101/2024.01.03.24300765

**Authors:** Xianghong Hu, Mingxuan Cai, Jiashun Xiao, Xiaomeng Wan, Zhiwei Wang, Hongyu Zhao, Can Yang

## Abstract

Mendelian Randomization (MR), which utilizes genetic variants as instrumental variables (IVs), has gained popularity as a method for causal inference between phenotypes using genetic data. While efforts have been made to relax IV assumptions and develop new methods for causal inference in the presence of invalid IVs due to confounding, the reliability of MR methods in real-world applications remains uncertain. To bridge this gap, we conducted a benchmark study evaluating 15 MR methods using real-world genetic datasets. Our study focused on three crucial aspects: type I error control in the presence of various confounding scenarios (e.g., population stratification, pleiotropy, and assortative mating), the accuracy of causal effect estimates, replicability and power. By comprehensively evaluating the performance of compared methods over one thousand pairs of exposure-outcome traits, our study not only provides valuable insights into the performance and limitations of the compared methods but also offers practical guidance for researchers to choose appropriate MR methods for causal inference.

## Introduction

Understanding the causal relationships between exposures and outcomes is crucial in biomedical and social science research, as it enables discoveries in etiology, aids in drug development, and informs policy-making. While randomized controlled trials (RCTs) are considered the gold standard for assessing causality, they can be time-consuming, costly, and sometimes ethically challenging [1]. Causal inference based on observational data presents its own challenges, such as unmeasured confounding or reverse causality. Mendelian randomization (MR) offers a promising approach to performing causal inference using observed genetic data [2, 3]. According to Mendel’s law of inheritance, genotypes are randomly inherited from parents to offspring, thereby ideally being independent of environmental confounding factors. This characteristic motivates researchers to explore genetic data in order to study the causal effects of one phenotype (exposure) on another phenotype (outcome). In recent years, MR has gained popularity due to the availability of summary statistics from thousands of genome-wide association studies (GWAS) covering a wide range of phenotypes. Leveraging the rich genetic data resources available, researchers worldwide can investigate the potential causal relationships between exposures and outcomes of interest, encompassing diverse applications such as identifying disease risk causation [4], providing evidence for epidemiological associations [5], and prioritizing targets in drug development [6, 7].

To perform causal inference using MR approaches, genetic variants (typically Single Nucleotide Polymorphisms, i.e., SNPs) serve as instrument variables (IVs). A valid IV should satisfy the following three IV assumptions [8, 9]: (1) it is associated with the exposure of interest; (2) it is not associated with the confounders of the exposure and outcome traits; and (3) it affects the outcome only through the exposure of interest. However, these assumptions underlying MR are often too strong to be satisfied in real applications. In recent years, much effort has been devoted to relaxing these assumptions and new MR methods have been designed to enable causal inference in the presence of invalid IVs. To name a few, MR-PRESSO [10], cML-MA [11], and MR-Lasso [12] use outlier detection to identify invalid IVs and remove them from the MR analysis. MR-Robust[12], weighted-median [13], and weighted-mode [14] use outlier-robust techniques to mitigate the effects of invalid IVs. Additionally, methods like Egger [15], RAPS [16], and BWMR [17] employ probabilistic models to correct for different types of pleiotropy, while CAUSE [18], MRAPSS [19], MRMix [20], MR-ConMix [21], and MR-CUE [22] employ mixture component models to characterize valid and invalid signals, enabling causal inference based on the component of valid signals.

Although considerable progress has been made in the development of MR methods, their robustness to the violation of underlying assumptions in real-world applications remains largely unclear. Due to the complexity of human genetics, several factors can significantly impact the performance of existing MR methods. Firstly, complex traits often exhibit high polygenicity, meaning that individual SNPs have small effect sizes. To satisfy the IV assumption (1), researchers select SNPs as IVs from the exposure GWAS using a *p*-value threshold (IV threshold). However, this selection process may inadvertently include weakly associated SNPs, which can introduce bias into MR estimates. Moreover, using the same exposure dataset for IV selection and MR estimation in two-sample MR settings can induce non-ignorable bias, known as selection bias [16]. Second, population stratification and family-level confounders (e.g., assortative mating and dynastic effects) are well-known issues in population-based GWAS, which can introduce associations between genetic instruments and unobserved confounders [23, 24, 25], leading to the violation of IV assumption (2). Despite the significance of population stratification and family-level confounders, many existing MR methods have not explicitly accounted for these. Third, pleiotropy is a ubiquitous phenomenon in human genetics, referring to a single genetic variant influencing multiple traits, thereby violating IV assumption (3) [26]. Carefully accounting for pleiotropy is crucial for reliable causal inference using MR approaches. Given these complexities, it is crucial to conduct benchmarking studies to assess the reliability of existing MR methods when their model assumptions may be violated. Such studies would provide valuable insights into the performance and limitations of these methods in real-world scenarios.

In this study, we present a benchmarking analysis of MR methods for causal inference with real-world genetic datasets. Our focus is on MR methods that utilize GWAS summary statistics as input, as they do not require access to individual-level GWAS data and are widely applicable. Specifically, we consider 15 MR methods, including the standard IVW (fixed) [27] and IVW (random) [28] and 13 other advanced MR methods: Egger, RAPS, Weighted-median, Weighted-mode, MR-PRESSO, MRMix, cML-MA, MR-Robust, MR-Lasso, MR-CUE, CAUSE, MRAPSS and MR-ConMix. To assess the performance of these MR methods, we utilized real-world datasets and focused on three key aspects: type I error control, the accuracy of causal effect estimates, and replicability. Particularly, in evaluating type I error control, we used GWAS summary-level datasets for over one thousand exposure-outcome trait pairs of no causal effect, serving as negative controls. These trait pairs were carefully selected to represent scenarios involving confounding factors, such as population stratification and pleiotropy. We conducted a comparison between population-based MR and family-based MR to evaluate the influence of family-level confounders. Through our comprehensive experiments using real-world datasets, we found that the performance of MR methods is heavily influenced by confounding factors that arise from various sources in practical scenarios. We also investigated the influence of summary-level data pre-processing steps, such as the inclusion of SNPs with different minor allele frequencies and the choice of reference genome panels. Our study offers practical guidelines for researchers in choosing appropriate MR methods and improving the reliability of causal inference in MR analyses.

## Results

### The experimental design for benchmarking MR methods

We conducted a benchmarking of 15 summary-level data-based MR methods, which were categorized into four groups: IVW-class, outlier detection and removal methods, model-based methods, and outlier robust methods (Fig. 1-A, Table S1, section 1 of the supplementary note). The procedure for running the MR methods is outlined in Fig. 1-B and described in detail in the Method section. To ensure a comprehensive evaluation, we utilized real-world datasets and focused on three crucial aspects: type I error control, the accuracy of causal effect estimates, replicability and power (see Fig. 1-C).

**Figure 1:**
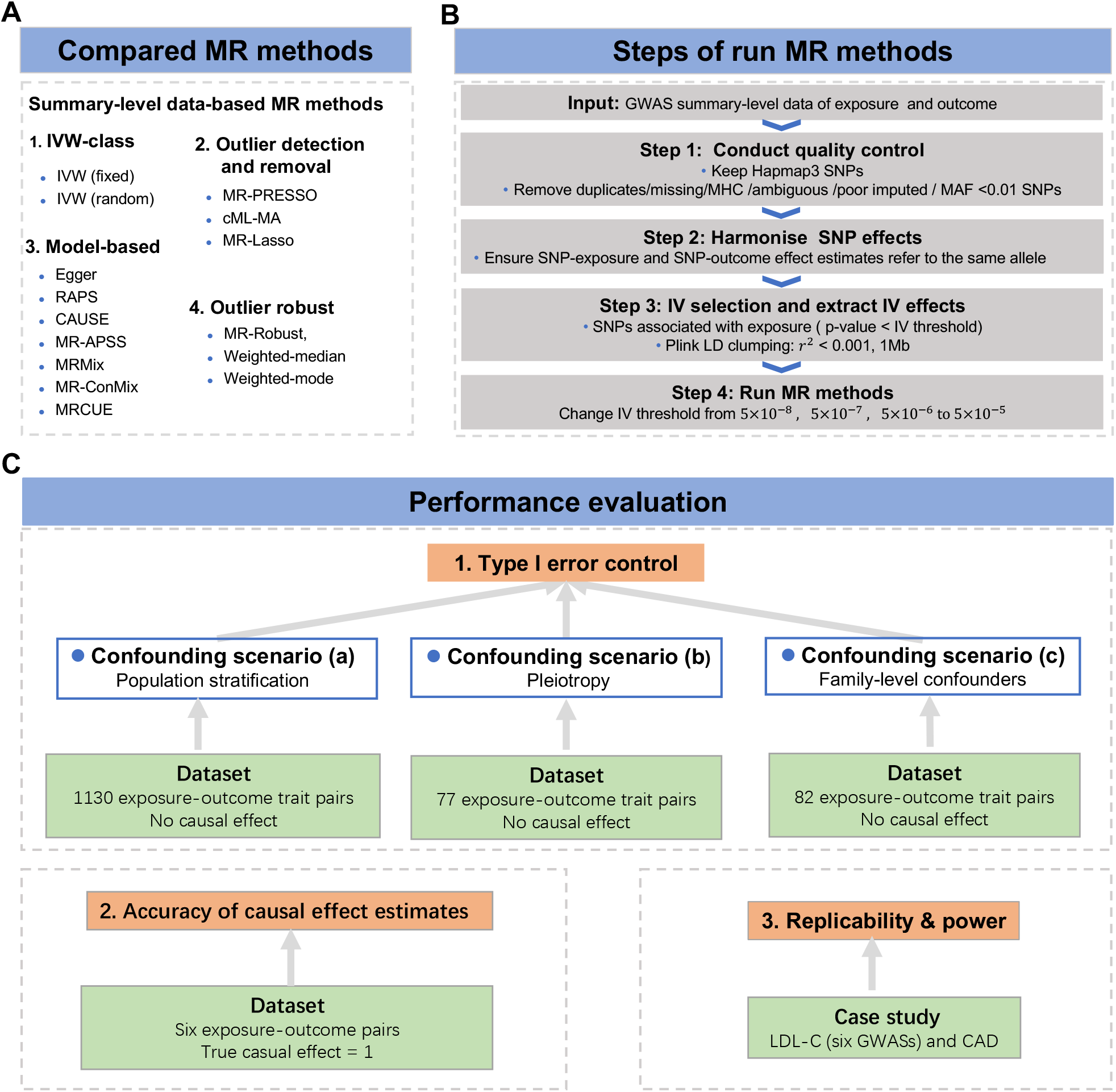
Experimental design for benchmarking MR methods. A We compared the performance of 15GWAS summary-level data-based MR Methods. B We designed a four-step procedure for running MR methods. C We used real-world datasets to evaluate the performance of MR methods on three aspects: Type I error control in three confounding scenarios, including (a) population stratification, (b) pleiotropy, and (c) family-level confounders, the accuracy of causal effect estimates, replicability and power.

To assess type I error control, we applied the MR methods to GWAS summary statistics for three sets of exposure-outcome trait pairs with no causal effect. The three sets of trait pairs represented three different confounding scenarios, including (a) population stratification, (b) pleiotropy, and (c) family-level confounders. Specifically, in scenario (a), we used 1,130 trait pairs between 226 exposures from UK Biobank and five negative control outcomes to investigate the influence of population stratification on MR methods (Supplementary data 1). The negative control study was designed carefully based on two criteria: First, the outcomes should not be causally affected by the exposures. Second, both the outcomes and exposures should be affected by population stratification. In this scenario, we chose four hair color-related traits and tanning ability as negative control outcomes. These traits are mainly determined at birth and are likely influenced by population stratification [24]. In scenario (b), we analyzed trait pairs between 11 exposures and seven negative control outcomes. The selected exposures included five adult behavior-related traits and six aging-related traits, while the negative control outcomes were seven childhood-related traits (Supplementary data 2). This choice was based on the convention that traits developed after adulthood are unlikely to affect traits developed before adulthood causally. These negative control outcomes exhibited non-zero genetic correlations with most of the exposures (SFig S4), indicating that pleiotropy is a major confounder here. In scenario (c), we analyzed 82 trait pairs using both population-based GWASs and family-based GWASs to examine the influence of family-level confounders (Supplementary data 3). Population-based GWAS estimates, which are derived from unrelated individuals, are known to be susceptible to bias due to the influence of family-level confounders. Conversely, family-based GWAS designs offer the advantage of accounting for the effects of family-level confounders when estimating GWAS effects [29, 30]. By comparing the results of MR analyses obtained from the population-based GWAS and family-based GWAS designs, we can assess the effectiveness of MR methods in controlling for type I errors in the presence of family-level confounding, such as assortative mating and dynastic effects. For this scenario, we required the trait pairs to be genetically uncorrelated. Based on the principle “no correlation implies no causal relationship”, we treated these trait pairs as negative controls. By applying MR methods to the three datasets representing different confounding scenarios, we investigated their ability to control type I errors in the presence of different confounding factors.

In evaluating the accuracy of causal effect estimates, we examined six pairs of traits where each pair comprised the same trait as both the “exposure” and the “outcome” (Supplementary data 4). The UK Biobank dataset was divided equally to obtain exposure and outcome GWAS data. Importantly, the true causal effects in this analysis were known to be exactly one [16, 31]. This design allowed us to assess the accuracy of MR methods in estimating causal effects. To evaluate replicability and power, we focused on a positive control example involving low-density lipoprotein cholesterol (LDL-C) and coronary artery disease (CAD). We applied all the MR methods to six GWAS datasets for LDL-C obtained from five distinct studies (Supplementary data 5). By analyzing multiple GWAS datasets for the same trait, we assessed the replicability of the causal effect estimates across different study designs and sample sizes. Detailed information about the datasets used in this study can be found in the Method section, and specific details regarding the sources of GWAS data are summarized in Stable 1-5.

Throughout the evaluation process, our first step was to assess the performance of MR methods when IVs were selected based on the default *p*-value thresholds in the exposure GWAS. Specifically, among the compared methods, MR-APSS and MR-CUE utilized a default IV threshold of 5 *×* 10*^−^*^5^, CAUSE employed a default IV threshold of 1 *×* 10*^−^*^3^, and the remaining methods required strong IVs with a default IV threshold of 5 *×* 10*^−^*^8^. All of the compared MR methods, except for MR-CUE, require Plink LD clump (*r*^2^ = 0.001,1Mb) to obtain independent SNPs as IVs. Furthermore, we introduced variations in the *p*-value thresholds used for IV selection to examine the methods’ performance across a range of IV thresholds, including a stringent threshold of 5 *×* 10*^−^*^8^, as well as more relaxed thresholds of 5 *×* 10*^−^*^7^, 5 *×* 10*^−^*^6^, and 5 *×* 10*^−^*^5^. As the IV thresholds become looser, the number of IVs, including both valid IVs and invalid IVs, may increase. Moreover, the number of IVs with weaker effects may also increase. This analysis allowed us to assess the robustness of these methods in handling invalid IVs due to confounding factors and determine whether they are sensitive to the choice of IVs used in MR analysis.

### MR-APSS, Egger, Weighted-mode, and CAUSE achieve better performance in type I error control

We conducted a comprehensive evaluation to assess the effectiveness of 15 MR methods in controlling type I errors across various confounding scenarios. The evaluation utilized three real-world datasets and focused on three specific scenarios: population stratification, pleiotropy, and family-level confounders. To evaluate the performance of these methods, we generated QQ plots to visualize the *p*-values produced by each method for the three datasets, as shown in Figs. 2-4. These QQ plots provide a visual tool to identify deviations from the expected diagonal line, which helps determine if the methods are generating systematically inflated or deflated *p*-values. In this analysis, we initially assessed the MR methods using their default setting for IV selection. Specifically, we first examined the performance of MR-APSS and MR-CUE at the IV threshold of 5 *×* 10*^−^*^5^, the performance of CAUSE at the IV threshold of 1 *×* 10*^−^*^3^, and the performance of other methods at the IV threshold of 5 *×* 10*^−^*^8^.

**Figure 2:**
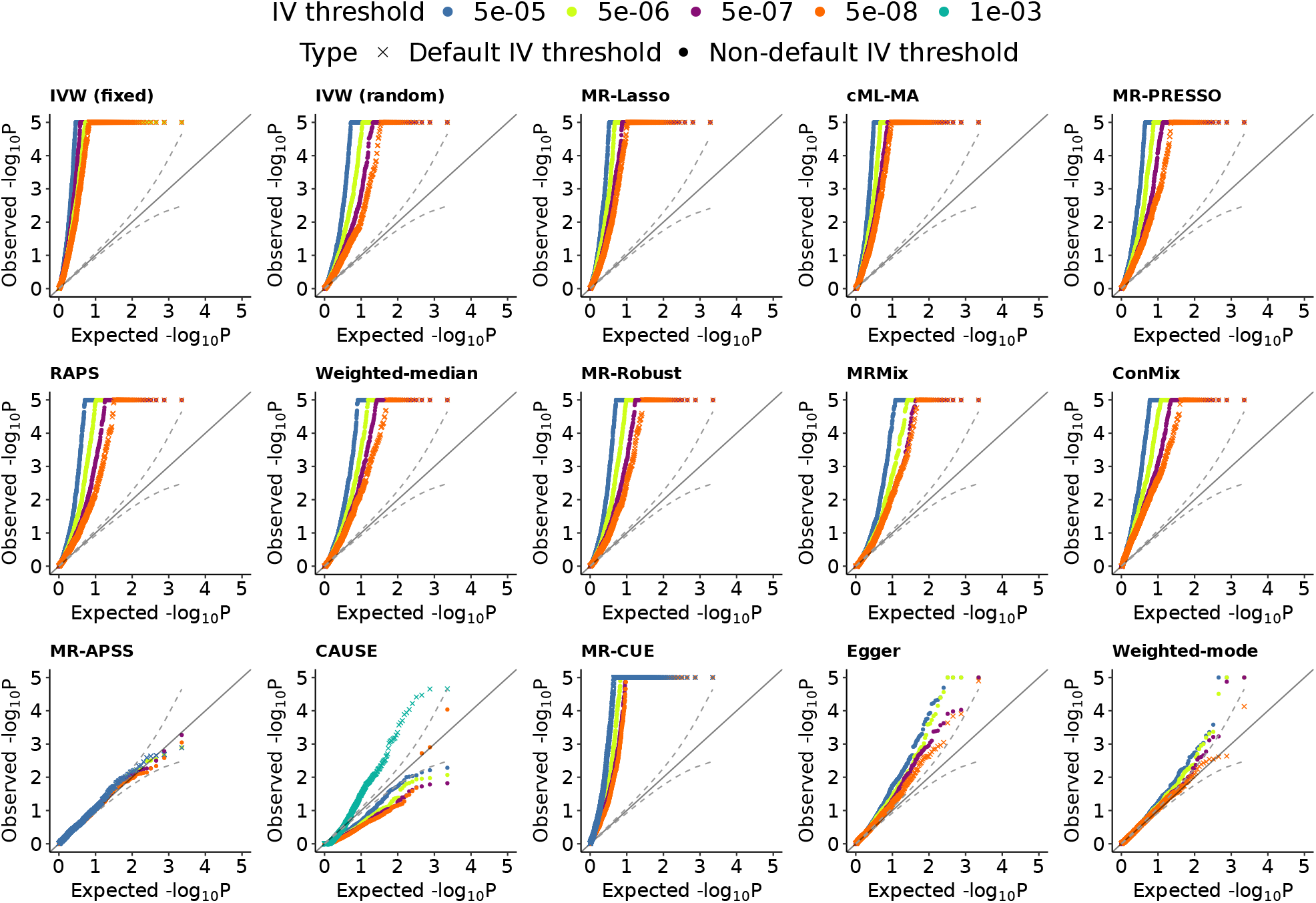
Evaluation of type I error control in confounding scenario (a) of population stratification. Type I error is evaluated by quantile-quantile plots of *−* log_10_(*p*) values from the 15 compared methods when testing the causal effect for 1130 negative control trait pairs at different IV thresholds. The 15 compared methods include IVW (fixed), IVW (random), Egger, RAPS, Weighted-median, Weighted-mode, MR-PRESSO, MRMix, cML-MA, MR-Robust, MR-Lasso, MR-CUE, CAUSE, MRAPSS and MR-ConMix. Each distinct color on the plot represents the results at a specific IV threshold and the results at the default IV thresholds of the compared MR methods are marked by a cross symbol. The default IV thresholds for MR-APSS and MR-CUE were set at 5 *×* 10*^−^*^5^, while the default IV threshold for CAUSE was set at 1 *×* 10*^−^*^3^. The remaining methods utilized a default IV threshold of 5 *×* 10*^−^*^8^.

In scenario (a), characterized by the presence of strong population stratification, MR-APSS and Weighted-mode consistently generated well-calibrated *p*-values, using their default IV thresholds. However, Egger’s *p*-values were slightly inflated. The p-values of CAUSE initially showed deflation but later exhibited inflation. Further analysis of causal effect estimates reveals that CAUSE’s confidence intervals are more reliable compared to its *p*-values. On the other hand, the remaining 11 methods, including IVW (fixed), IVW (random), RAPS, Weighted-median, MR-PRESSO, MRMix, cML-MA, MR-Robust, MR-Lasso, MR-CUE, and MR-ConMix, exhibited highly inflated *p*-values at the default IV threshold of 5 *×* 10*^−^*^8^. Notably, IVW (fixed) demonstrated the most severe inflation, which is expected as it is a basic MR method that does not account for IV invalidity, leading to bias and inflation in the estimates. While other methods incorporated different assumptions to address invalid IVs, they still failed to effectively control type I error inflation. These findings highlight the limitations of existing methods in handling scenarios involving strong population stratification, where their model assumptions do not align well with real-world situations.

In scenario (b), where pleiotropy is present, several methods exhibited effective control of type I errors. Notably, CAUSE, Egger, MR-APSS, and Weighted-mode demonstrated the absence of inflated *p*-values, indicating their capability to address pleiotropy. However, it was observed that CAUSE’s *p*-values were deflated. On the other hand, several other methods, including IVW (random), MR-Lasso, MR-PRESSO, RAPS, Weighted-median, IVW (fixed), cML-MA, MR-ConMix, and MR-Robust, exhibited inflated *p*-values at the default IV threshold.

Notably, IVW (fixed), cML-MA, and MR-ConMix showed more pronounced inflation compared to the other methods. Despite these methods’ primary focus on addressing pleiotropy, their performance in controlling type I errors was not entirely satisfactory. This observation indicates the ongoing challenge in effectively handling pleiotropy in MR analysis and the need for further methodological advancements.

In scenario (c), we conducted a comparison between the results of MR methods using both population-based and family-based GWAS data for 82 negative control trait pairs. The objective was to assess the effectiveness of MR methods in controlling for type I errors in the presence of family-level confounders. The QQ plots for MR methods at their default IV thresholds, using both population-based GWAS and within-family-based GWAS summary-level data, are depicted in Fig. 4. When using population-based GWAS data, Egger, Weighted-mode, and MRMix did not yield inflated *p*-values. CAUSE produced deflated *p*-values, and MR-APSS exhibited very slight inflation in the *p*-values. On the other hand, other methods such as IVW (fixed), MR-Lasso, cML-MA, and MR-CUE produced inflated *p*-values, indicating challenges in adequately addressing family-level confounding using these methods. However, when utilizing family-based GWAS data, all MR methods produced well-calibrated *p*-values, demonstrating effective control of type I error inflation. Our results provide further evidence for the usefulness of family-based MR in mitigating the influence of family-level confounders in MR analysis.

**Figure 3:**
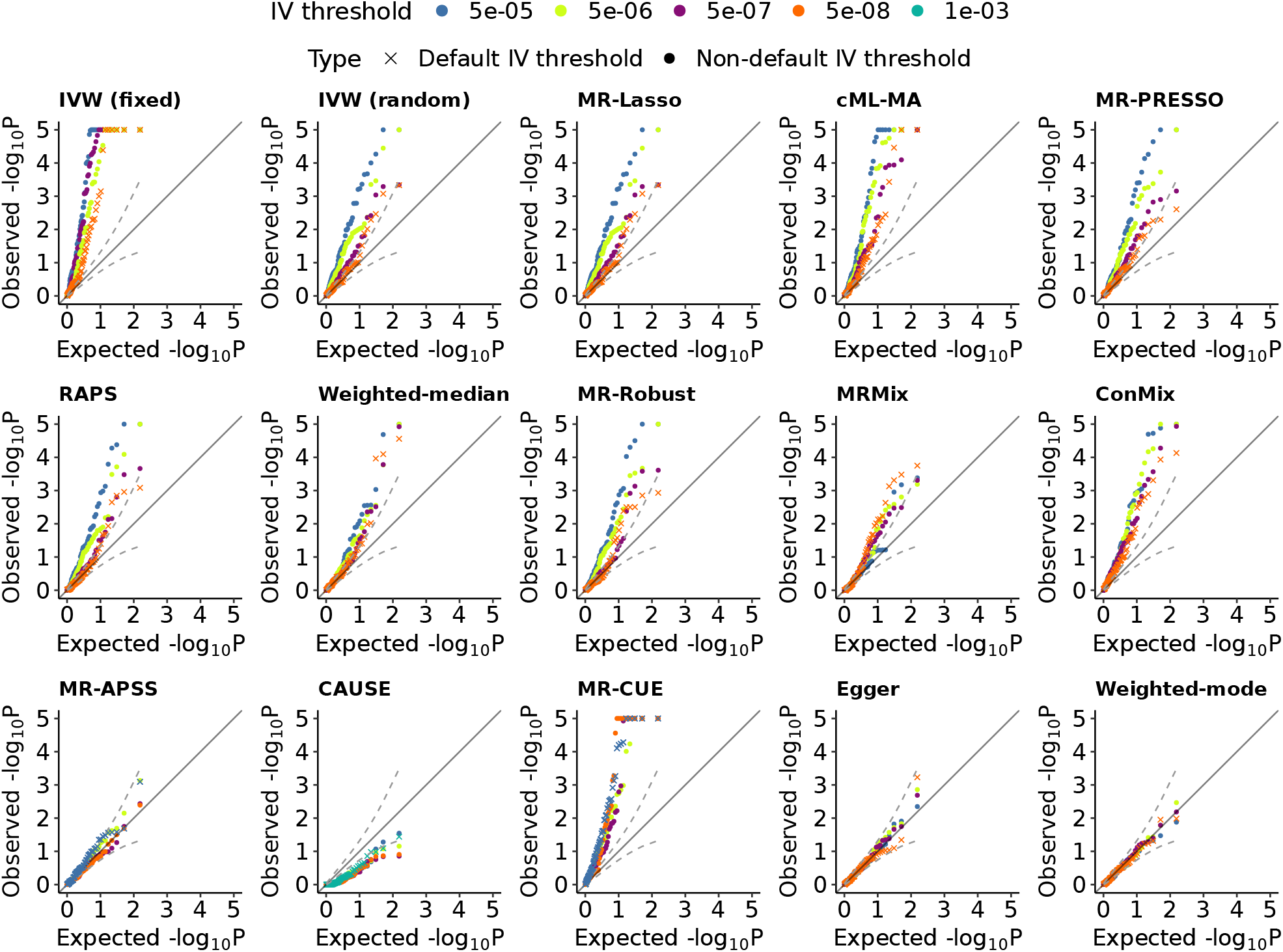
Evaluation of type I error control in confounding scenario (b) of pleiotropy. Type I error is evaluated by quantile-quantile plots of *−* log_10_(*p*) values from the 15 compared methods when testing the causal effect for 77 negative control trait pairs at different IV thresholds. The 15 compared methods include IVW (fixed), IVW (random), Egger, RAPS, Weighted-median, Weighted-mode, MR-PRESSO, MRMix, cML-MA, MR-Robust, MR-Lasso, MR-CUE, CAUSE, MRAPSS and MR-ConMix. Each distinct color on the plot represents the results at a specific IV threshold and the results at the default IV thresholds of the compared MR methods are marked by a cross symbol. The default IV thresholds for MR-APSS and MR-CUE were set at 5 *×* 10*^−^*^5^, while the default IV threshold for CAUSE was set at 1 *×* 10*^−^*^3^. The remaining methods utilized a default IV threshold of 5 *×* 10*^−^*^8^.

**Figure 4:**
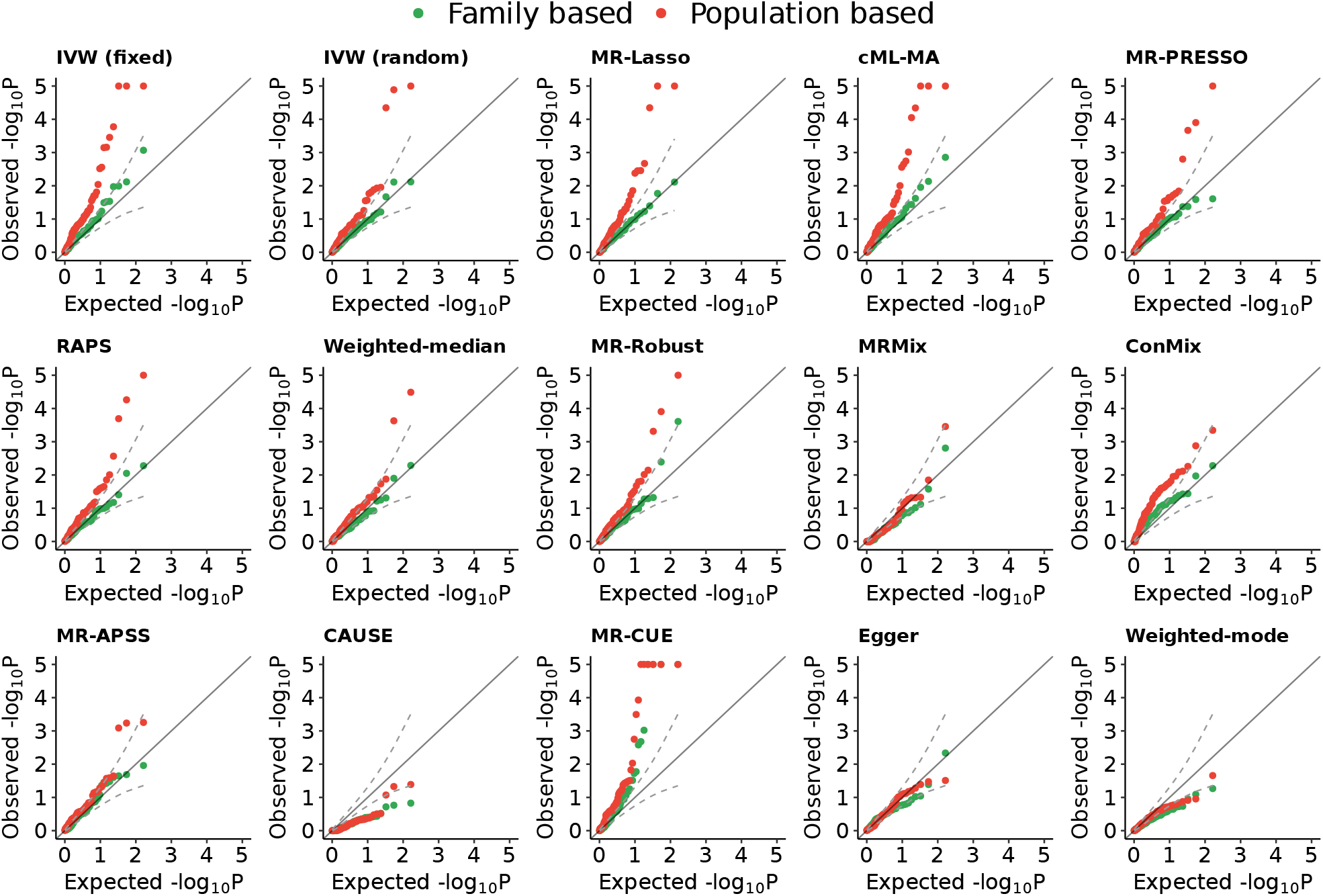
Evaluation of Type I error control in the confounding scenario (c) of family-level confounders. Quantile-quantile (Q-Q) plots illustrating the *−* log_10_(*p*) values for testing causal effects on 82 trait pairs using 15 different methods at their default IV thresholds. The comparison includes results from both population-based GWASs (depicted as red triangles) and sibling-based GWASs (depicted as green dots). The evaluated methods consist of IVW-fixed, IVW-random, Egger, RAPS, Weighted-median, Weighted-mode, MR-PRESSO, MRMix, cML-MA, MR-Robust, MR-Lasso, MR-CUE, CAUSE, MRAPSS, and MR-ConMix. MR-APSS and MR-CUE employ an IV threshold of 5 *×* 10*^−^*^5^, CAUSE uses a threshold of 1 *×* 10*^−^*^3^, while the remaining methods use a threshold of 5 *×* 10*^−^*^8^.

### IV selection largely affects the performance of MR methods

We conducted a comprehensive investigation into the performance of various MR methods by analyzing their behavior across a range of IV thresholds, including a stringent threshold of 5 *×* 10*^−^*^8^, as well as more relaxed thresholds of 5 *×* 10*^−^*^7^, 5 *×* 10*^−^*^6^, and 5 *×* 10*^−^*^5^. The QQ plots in Figs. 2 and 3 depict the results obtained in confounding scenarios (a) and (b), respectively. From these plots, we can observe that MR-APSS, Egger, and weighted-mode consistently generated well-calibrated *p*-values across varying IV thresholds. However, it is worth noting that the *p*-values obtained from CAUSE were consistently deflated. On the other hand, the remaining 11 methods, including IVW (fixed), IVW (random), RAPS, Weighted-median, MR-PRESSO, MRMix, cML-MA, MR-Robust, MR-Lasso, MR-CUE, and MR-ConMix, exhibited substantially inflated *p*-values. Furthermore, the degree of *p*-value inflation tended to increase as the IV threshold became looser. MRMix was an exception with slightly inflated *p*-values at a less stringent IV threshold of 5 *×* 10*^−^*^5^ but more inflated *p*-values at the IV threshold of 5 *×* 10*^−^*^8^, as observed in Fig. 3. This observation suggests that MRMix can be sensitive to the number of IVs used. It tends to produce more false positives when there is a limited number of IVs. Our results indicate that causal inference results obtained from most of the methods are sensitive to the IV threshold.

### Accuracy of causal effect estimates

To assess the accuracy of MR methods in estimating causal effects, we examined six pairs of traits, where each pair involved the same trait being considered as both the “exposure” and the “outcome” [16, 31]. In this specific scenario, the true causal effects for these trait pairs were precisely known to be equal to one. This knowledge enabled us to compare the accuracy of causal effect estimates given by different MR methods. We included three continuous traits: Height, Waist Circumference (WC), and Educational Attainment (EA), as well as three binary traits: Hypertension, High cholesterol, and Asthma. For each trait, we divided the UK Biobank samples into two halves, representing the exposure GWAS and the outcome GWAS. We utilized the Bolt-LMM software [32] to obtain GWAS summary statistics from these subsets. The exposure GWAS summary statistics were used for both IV selection and causal effect estimation.

We varied the IV thresholds, starting with a stringent threshold of 5 *×* 10*^−^*^8^, and progressively relaxed the thresholds to 5 *×* 10*^−^*^7^, 5 *×* 10*^−^*^6^, and 5 *×* 10*^−^*^5^. As such, we can investigate the robustness of MR methods to weak IV bias and selection bias. However, it is important to note that in this analysis, we cannot assess the robustness of MR methods to pleiotropy or other forms of confounding when testing the effect of the trait on itself using data from the same population. The causal effect estimates and their confidence intervals for 15 MR methods at different IV thresholds are presented in Fig. 5.

**Figure 5:**
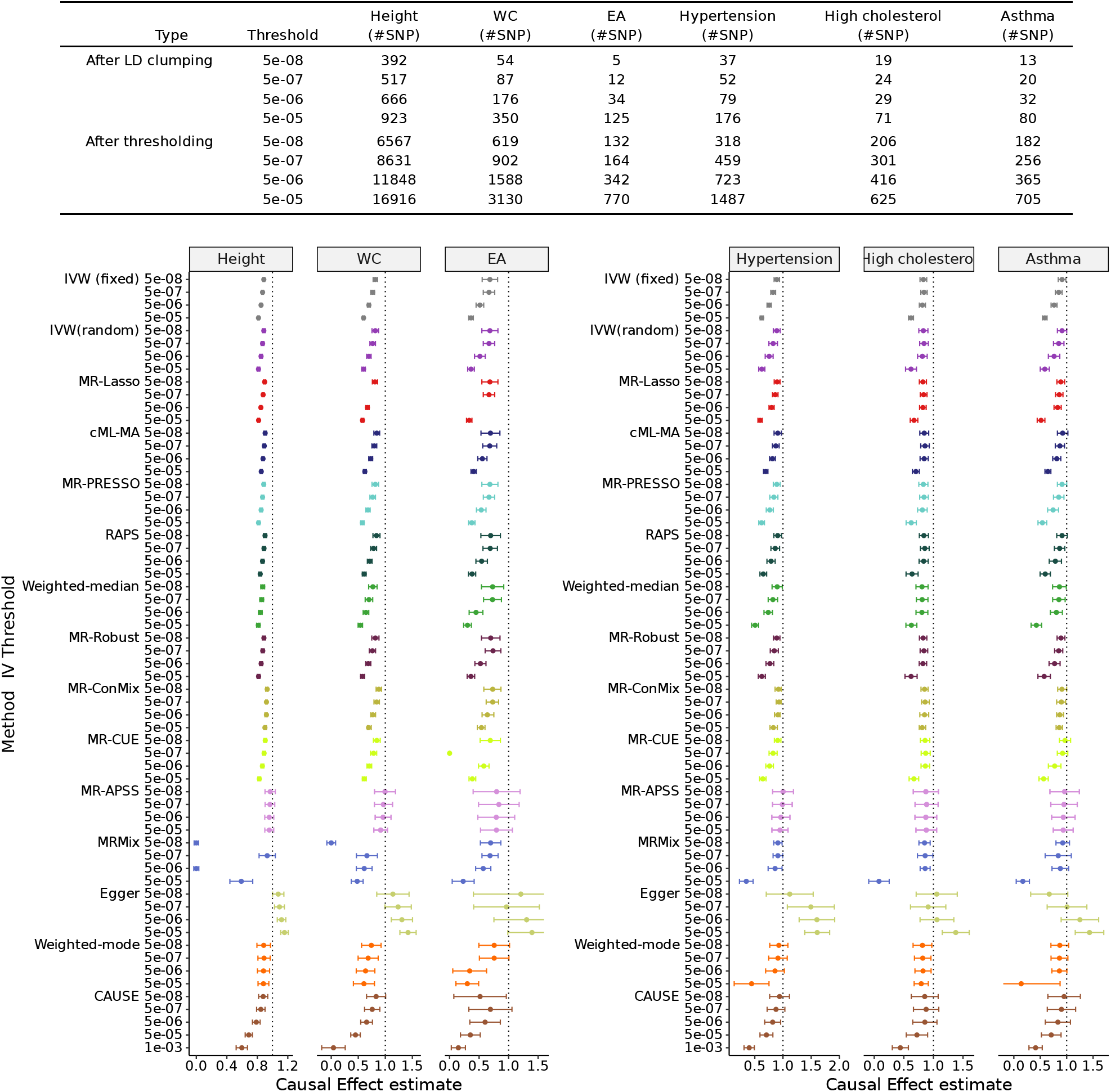
Evaluation of the accuracy of causal effect estimation of 15 MR methods for six trait pairs. Each pair comprised the same trait as both the “exposure” and the “outcome” Analyzed traits include three continuous traits, i.e., Height, Waist Circumference (WC), and Educational Attainment (EA), and three binary traits, i.e., Hypertension, High cholesterol, and Asthma. The top panel shows the number of IVs selected using different IV thresholds with/without LD clumping. The bottom panel shows the point estimates and 95% confidence intervals of different methods at different IV thresholds for the three continuous traits (bottom left) and the three binary traits (bottom right). The vertical dashed gray line represents the true causal effect size 1. Each of the 15 MR methods is represented by a distinct color.

Our study found that MR-APSS outperformed other MR methods and produced more accurate causal effect estimates that were closer to the true value. Importantly, all of the confidence intervals produced by MR-APSS at different IV thresholds covered the true value. This indicates that MR-APSS is a promising method for accurately estimating causal effects in MR analyses, robust to weak IV bias and selection bias. Furthermore, MR-APSS produces narrower confidence intervals as the IV selection threshold was relaxed and weaker IVs were included in the MR analysis. These findings highlight the potential advantages of including more weak IVs in MR analysis to increase statistical power. Weighted-mode, at its default IV threshold of 5 *×* 10*^−^*^8^, delivered estimates comparable to those of MR-APSS in terms of accuracy and coverage of true causal effects within the confidence intervals. Egger, while producing larger estimation errors, provided unbiased estimates when a stringent IV threshold was applied (5 *×* 10*^−^*^8^). However, it tended to overestimate causal effects when a looser IV threshold was used. CAUSE, on the other hand, produced confidence intervals covering the true causal effect only at a stringent threshold of 5 *×* 10*^−^*^8^.

The majority of existing MR methods, including IVW (fixed), IVW (random), MR-Lasso, cML-MA, MR-PRESSO, RAPS, Weighted-median, MR-Robust, MRMix, and MR-ConMix, displayed limitations in estimation accuracy in the presence of weak IV bias and selection bias. As the IV threshold became looser and weaker instruments were included, these methods produced estimates that were biased toward the null effect. Moreover, their confidence intervals failed to cover the true causal effects in most cases. This indicates that these methods are not capable of dealing with weak IV bias and selection bias, which compromises the accuracy of the causal effect estimates. It is crucial to acknowledge that the current strategy of using a stringent IV threshold for IV selection, such as 5 *×* 10*^−^*^8^, is not a foolproof solution to address weak IV bias and selection bias. This approach has its limitations, including reduced power due to a limited number of IVs and susceptibility to weak IV bias and selection bias even with a stringent threshold. Our findings highlight the need for more robust MR methods that can effectively handle weak instruments and mitigate selection bias to accurately estimate causal effects.

In addition to biased causal effect estimation, methods such as MRMix and MR-Lasso have their specific limitations. MRMix exhibited some instability when varying IV thresholds. For instance, when examining the effect of height on itself, MRMix estimated the causal effect as 0 with a standard error of 0.015 using 302 IVs at a threshold of 5 *×* 10*^−^*^8^. Similarly, at an IV threshold of 5 *×* 10*^−^*^6^ with 666 IVs, MRMix again estimated the causal effect as 0 with a standard error of 0.017. However, the estimation result by MRMix at an IV threshold of 5 *×* 10*^−^*^7^ was much more reliable. In this case, the causal effect of height on itself was estimated as 0.93 with a standard error of 0.055, utilizing 517 IVs. MR-Lasso failed to report causal estimates in some cases. For example, when testing the causal effect of WC on itself at the IV threshold of 5 *×* 10*^−^*^7^ and that of EA on itself at the IV threshold of 5 *×* 10*^−^*^6^, MR-Lasso detected all IVs as invalid outliers and did not report any causal estimates.

### Replicability and power

To assess the replicability and power of the MR methods, we applied all the compared methods to infer the causal effect between LDL-C and CAD using six LDL-C GWAS datasets collected from five separate studies. By comparing the causal effects estimated from different GWAS datasets of the same trait, we were able to assess the reliability and generalizability of their causal effect estimates across different study designs and sample sizes. We also considered the IV thresholds varied from the stringent 5 *×* 10*^−^*^8^ to the relaxed 5 *×* 10*^−^*^7^, 5 *×* 10*^−^*^6^, and 5 *×* 10*^−^*^5^ to examine the sensitivity of the methods to different levels of instrument strength. The causal effect estimates, their 95% confidence intervals, and *p*-values produced by each method for different datasets and different IV thresholds are shown in Fig. 6.

**Figure 6:**
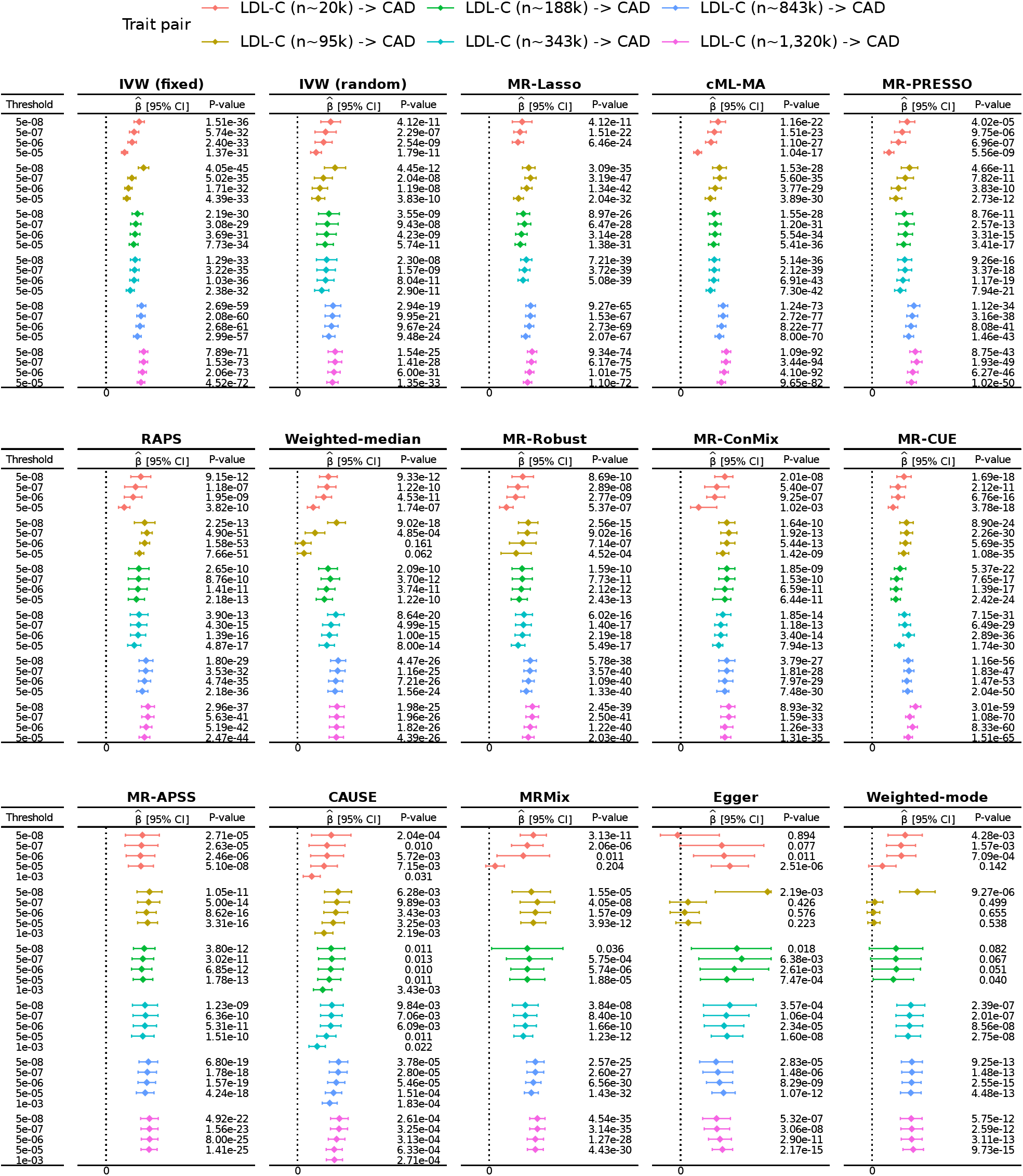
MR analysis results of LDL-C and CAD from 15 MR methods at different IV thresholds. Six GWAS datasets for LDL-C were used, each represented by a distinct color. The vertical dashed gray line represents the zero causal effect size. Compared methods include IVW-fixed, IVW-random, Egger, RAPS, Weighted-median, Weighted-mode, MR-PRESSO, MRMix, cML-MA, MR-Robust, MR-Lasso, MR-CUE, CAUSE, MRAPSS and MR-ConMix represented by different colors. The IV threshold is varied from 5 *×* 10*^−^*^5^, 5 *×* 10*^−^*^6^, 5 *×* 10*^−^*^7^ and 5 *×* 10*^−^*^8^. The result of CAUSE at its default IV threshold 1 *×* 10*^−^*^3^ is also presented.

Among all the compared methods, CAUSE and MR-APSS achieved outstanding performance in terms of replicability. Both CAUSE and MR-APSS are capable of producing confidence intervals that reject the null causal effect. The causal effect estimates and confidence intervals produced by both methods were highly consistent across different studies and different IV thresholds. The high consistency and generalizability of the results produced by these methods are particularly noteworthy, as they suggest that the causal effect estimates obtained using CAUSE and MR-APSS are likely to be more accurate and reliable than those obtained using other methods. However, we note that the *p*-values produced by CAUSE do not agree well with its confidence intervals. Consistent with previous results, the *p*-values produced by CAUSE are likely to be deflated. Therefore, caution should be exercised when interpreting the *p*-values produced by CAUSE.

Most of the MR methods we compared detected a significant causal relationship for all 24 tests between LDL-C and CAD using six datasets of LDL-C at four different IV thresholds. However, Egger, Weighted-mode, Weighted-median, and MRMix were unable to detect significant causal relationships in some cases. Specifically, among the 24 tests, Egger, Weighted-mode, Weighted-median, and MRMix failed to detect significant causal effects for five, four, two, and one test between LDL-C and CAD at the nominal level of 0.05, respectively. Although Egger and Weighted-mode showed good performance in terms of type I error control, our analysis revealed that Egger tended to produce estimates with large estimation errors, and Weighted-mode may have low power. Moreover, Weighted-median and MRMix, which are likely to produce false positives as shown in our previous analysis, can also lead to causal effects being wrongly shrunk to zero in some cases. However, we found that all methods, including Egger, Weighted-mode, Weighted-median, and MRMix, were able to detect significant causal effects in the cases of LDL-C(*n ∼* 843k) and LDL-C(*n ∼* 1, 320k), indicating improved performance with large sample sizes. This suggests that larger sample sizes may lead to more accurate and reliable causal inference in MR analyses.

Consistent with previous findings, our analysis showed that most MR methods produced causal effect estimates that were sensitive to the choice of IV thresholds. Specifically, we found that causal effect estimates produced by most MR methods were closer to zero at a looser IV threshold of 5 *×* 10*^−^*^5^ compared to a more stringent IV threshold of 5 *×* 10*^−^*^8^. However, we observed better consistency in the causal estimates produced by the MR methods across IV thresholds for the cases of LDL-C(*n ∼* 843k) and LDL-C(*n ∼* 1, 320k). Importantly, as GWAS sample sizes increase to the scale of millions, the influence of weak IV bias and selection bias may be greatly alleviated. Our analysis highlights the potential benefits of larger sample sizes for improving the accuracy and reliability of MR analyses. Therefore, researchers should consider using larger sample sizes in MR studies to improve the robustness of their causal inference.

## Discussion

We present a benchmarking study of 15 two-sample summary-level data-based MR methods for causal inference. Our evaluation focuses on three crucial aspects: type I error control in the presence of various confounding scenarios (e.g., population stratification, pleiotropy, and assortative mating), the accuracy of causal effect estimates, replicability and power. Additionally, we explored the robustness of MR methods by evaluating their performance across a range of IV thresholds, assessing their ability to handle invalid IVs, and their sensitivity to IV selection. What sets our study apart is that our benchmark study is based on real-world datasets. Rather than relying on simulated or synthetic data, we carefully curated five diverse genetic datasets containing over one thousand trait pairs. The utilization of real-world datasets provides a more realistic and comprehensive evaluation of the performance of MR methods in practical scenarios.

Through the innovative designs of experiments that include a wide range of scenarios using real-world datasets, our study revealed that the performance of MR methods depends on underlying confounding factors that are very prevalent in real-world scenarios. Among the methods analyzed, Egger, weighted mode, and MR-APSS consistently demonstrated effective control of type I error across all three datasets representing different confounding scenarios, including population stratification, pleiotropy, and family-level confounders. However, CAUSE failed to control type I error using its default IV threshold of 1 *×* 10*^−^*^3^ in the presence of strong population stratification, although it exhibited deflation in other confounding scenarios. The remaining 11 MR methods displayed varying performances across the datasets representing different confounding scenarios. In the dataset representing strong population stratification (confounding scenario a), all 11 methods exhibited significant inflation of type I error. Conversely, in the datasets representing confounding scenarios of pleiotropy and family-level confounders (scenarios b and c), some methods, such as IVW (random), MR-Lasso, MR-PRESSO, RAPS, Weighted-median, IVW (fixed), cML-MA, MR-ConMix, and MR-Robust, demonstrated less severe inflation. Notably, MRMix displayed effective control of type I error in the dataset representing family-level confounders (confounding scenario c) but exhibited inflation in the dataset representing population stratification (confounding scenario a) and pleiotropy (confounding scenario b). These findings underscore the necessity of considering the characteristics of the datasets when selecting an appropriate MR method for analysis. Researchers should carefully assess the specific confounding factors present in their data and choose a method that has demonstrated robustness in handling those confounders.

Our study emphasized the significant impact of IV selection on the performance of MR methods. We found that using a looser threshold for IV selection resulted in inflated type I errors and increased bias in causal effect estimates for most methods. This highlights the limitations of certain MR methods in handling invalid or weak IVs and emphasizes the need to mitigate the potential bias associated with IV selection.

Based on our findings, we put forward the following recommendations as guidelines for best practices, aiming to assist researchers in choosing the most suitable summary-level MR methods for studying causal relationships between specific exposure-outcome trait pairs. By adhering to these guidelines, researchers can enhance the reliability and validity of their MR analyses. Firstly, we recommend conducting an analysis using negative controls. By incorporating negative controls, such as using hair colors as negative control outcomes, researchers can detect the presence of confounding bias and evaluate the robustness of different methods to confounding. This helps in selecting methods that can effectively handle confounding and provide more reliable results. Secondly, we advocate for adopting multiple standards for IV selection. Instead of relying solely on a single *p*-value threshold, researchers should consider various criteria and adjust the threshold accordingly to select IVs. By employing multiple standards, researchers can assess the sensitivity of MR methods to IV selection and invalid IVs. This allows for a more thorough evaluation of the methods’ performance and helps prioritize methods that are robust to IV selection. Lastly, whenever feasible, we encourage researchers to gather data from multiple independent sources for the exposure and outcome of interest. This could involve incorporating data from different study populations, cohorts, or databases. By considering data from diverse sources, researchers can prioritize methods that demonstrate high replicability across multiple sources. This increases the reliability of the findings and strengthens the credibility of the robustness of the selected MR method.

While our benchmark study provides valuable insights into the performance of summary-level MR methods, it does have certain limitations. Firstly, the selection and measurement of confounding factors in real-world datasets can be a challenging task. Although we made careful efforts to include datasets that represented specific confounding scenarios, it is important to recognize that different types of confounders may coexist in these datasets. Secondly, due to the difficulty in collecting true positive cases from real data, we assessed the estimation accuracy of causal effect by treating the same trait as both exposure and outcome and examined replicability with a case study by employing multiple GWASs of the same exposure trait. While these strategies indirectly reflect the performance of MR methods in terms of power, a more comprehensive power analysis using multiple positive cases would provide valuable insights into the methods’ ability to detect causal effects under different conditions. Thirdly, our evaluation of the estimation accuracy of MR methods utilized trait pairs where the exposure and outcome were the same trait. This design choice is currently the only possible way to ensure the true causal effects between trait pairs are known. However, we have to admit that the downside of this design is that this example does not test the methods’ robustness to confounding factors like pleiotropy because the exposure and outcome are the same traits. Lastly, our benchmark study focused solely on summary-level MR methods, but it is important to recognize the availability of individual-level MR methods such as GENIUS[33] and GENIUS-MAWII [34] and MR-MiSTERI [35]. Although these methods are beyond the scope of our study, researchers should consider exploring them when they align with the study design and data availability, as they may provide additional insights and benefits in specific research contexts.

## Methods

### Datasets for evaluation of type I error control in different confounding scenarios

#### Confounding scenario (a): Population stratification

We aim to assess the effectiveness of MR methods in controlling type I errors in the presence of population stratification. To achieve this, we chose four hair color-related traits (Hair color: black, Hair color: blonde, Hair color: light brown, Hair color: dark brown) and skin tanning ability (Tanning) as our negative control outcomes. The GWAS summary statistics for the negative control outcomes were obtained from the GWAS ATLAS resource [30], which contains GWAS data from 600 traits in the UK Biobank. These traits were selected based on their characteristics: they are primarily determined at birth and thus are unlikely to be influenced by traits occurring after birth, and they are susceptible to confounding due to population stratification as indicated by LDSC intercept values of 1.678 (se = 0.017) for Hair color: black, 1.206 (se = 0.016) for Hair color: blonde, 1.335 (se = 0.013) for Hair color: light brown, 1.510 (se = 0.018) for Hair color: dark brown, and 1.916 (se = 0.020) for Tanning.

Next, we focused on selecting suitable exposure traits from the remaining 555 traits available in the GWAS ATLAS. We applied specific criteria to identify traits that were unrelated to hair or skin, had LDSC heritability estimates greater than 0.01, and possessed a minimum of four IVs. Through this process, we identified 226 traits that met these criteria, which we then utilized as exposure traits in our MR analysis.

We then applied MR methods to the 1130 exposure-outcome trait pairs formed by the selected exposure and negative control outcome traits (Supplementary data 1) and evaluated the effectiveness of MR methods in controlling for type I errors in the presence of population stratification.

#### Confounding scenario (b): Pleiotropy

We aim to assess the type I error control of MR methods in the presence of confounding factors, such as pleiotropy, which can induce genetic correlation between trait pairs that are not causally linked. To accomplish this, we analyzed trait pairs consisting of 11 exposures and seven negative control outcomes. The selected exposures included five adult behavior-related traits, namely Coffee consumption [36], Instant coffee consumption [36], Ground coffee consumption [36], automobile speeding propensity [37] and risk [37], as well as six aging-related traits, including Self-rated health (http://www.nealelab.is/uk-biobank/, Phenotype Code: 2178), Longevity [38], Parental lifespan [39], Health span [40], Perceived age [41], and Frailty Index [42]. On the other hand, the negative control outcomes comprised seven childhood-related traits, such as Childhood aggression [43], Childhood BMI [44], Childhood intelligence [45], Fetal birth weight [46], Maternal birth weight [46], Pubertal growth (a single height measurement at age 10 in girls and 12 in boys) [47], and Comparative body size at age 10 [30]. We chose negative control outcomes based on the convention that traits developed after adulthood are unlikely to affect traits developed before adulthood causally. Consequently, causal effects between the selected exposures and negative control outcomes were considered implausible.

To conduct our analysis, we collected GWAS summary statistics for the exposure and outcome traits from multiple GWAS sources (detailed information can be found in Supplementary data 2). Subsequently, we examined the LDSC intercept estimates of the outcomes and calculated the genetic correlation estimates between the trait pairs (see SFig. S4). The LDSC intercepts for the outcomes were found to be approximately one, suggesting that population stratification was not a prominent confounding factor among these trait pairs. Out of the 77 trait pairs analyzed, 49 pairs exhibited significant genetic correlations at the nominal level of 0.05. These analyses allowed us to evaluate the performance of the MR methods in the presence of pleiotropy or other types of confounding that could induce genetic correlation between trait pairs. By considering these factors, we gained valuable insights into how well the MR methods controlled type I errors in the presence of confounders, thereby enhancing our understanding of their performance in such scenarios.

#### Confounding scenario (c): Family-level confounders

To assess the type I error control of MR methods in the presence of family-level confounders like assortative mating or other indirect genetic effects, we conducted an analysis using summary data obtained from a recent within-sibship GWAS study [30]. This dataset provided summary statistics for 25 traits, encompassing both within-sibship and population-based GWAS estimates. Details on these GWASs are summarized in Supplementary data 3. To ensure that the trait pairs analyzed in our study were suitable for evaluating type I errors, we required them to be genetically uncorrelated. This criterion was established to ensure that pairs with zero genetic correlation are unlikely to be causally linked, indicating the absence of a causal effect. To achieve this, we utilized LDSC [48] to estimate the genetic correlation between trait pairs among the 25 phenotypes using both population-based GWAS and within-sibship GWAS. Our selection process involved identifying 82 trait pairs (Supplementary data 3) with insignificant genetic correlation at the nominal level of 0.05 in both types of GWAS analyses. Subsequently, we applied MR methods to these selected trait pairs using both population-based GWAS and within-sibship GWAS. By comparing the results obtained from each method based on the two types of GWAS designs, we were able to examine the ability of MR methods to control for the effects of family-level confounders.

### Datasets for evaluation of the accuracy of causal effect estimates

To evaluate the accuracy of the causal effect estimates of each method, we consider a special setting where the exposure and outcome are the same traits. Under a linear model setting, the genetic effects of IVs on the exposure and the outcome are the same but the effect size estimates are different. Therefore, there is no pleiotropy or other forms of confounding, and we could expect the true causal effect known to be exactly one [16, 31, 49]. Specifically, we used six traits in this setting including three continuous traits, i.e. Height, Waist Circumference (WC), and Educational attainment (EA), and three binary traits, i.e. Hypertension, High cholesterol, and Asthma. To obtain the exposure GWAS and outcome GWAS, we split the UK Biobank samples into two halves. One half was used as the exposure GWAS and the other half was used as the outcome GWAS. The sample sizes of the GWASs ranged from 121,194 to 168,300 (see details in supplementary data 4). The GWAS summary statistics are obtained using the BOLT-LMM software [50]. In our analysis, GWAS estimates for the binary traits are also obtained using linear models through BOLT-LMM and are then used as input for MR analysis. We could thus expect the true causal effect between the same binary exposure and binary outcome also equal one.

### Datasets for evaluation of replicability and power

We used the example of low-density lipoprotein cholesterol (LDL-C) and coronary artery disease (CAD) for evaluation of the replicability and power of MR methods. The use of the LDL-C and CAD example in our case study provides several benefits. First, it serves as a positive control for comparing the performance of MR methods. High-level LDL-C is a well-established important risk factor for CAD. Several randomized control trials have consistently shown that lowering LDL-C levels with statins is effective in the prevention of CAD [51, 52, 53, 54, 55, 56, 57]. This allows us to evaluate the accuracy and replicability of different MR methods in a setting where we have high confidence in the existence of the positive causal effect. Second, the availability of multiple GWAS summary datasets for LDL-C provides a rich source of data for evaluating the performance of MR methods. We can thus assess the replicability of different MR methods using datasets with varying sample sizes and study designs. In our analysis, we gathered six European ancestries GWAS summary datasets for LDL-C i.e., LDL-C (*n ∼* 20k) [58], LDL-C (*n ∼* 95k) [59], LDL-C (*n ∼* 188k) [60], LDL-C (*n ∼* 343k) by the Neale Lab, LDL-C (*n ∼* 843k) (without UK biobank samples) and LDL-C(*n ∼* 1, 320k) (with UK biobank samples) [61]. The GWAS sample size increased from 19,840 in 2009 to 1.35 million in 2022. We used the same outcome GWAS for CAD which was obtained from the CARDIoGRAMplusC4D Consortium [62]. More details for the GWAS sources can be found in Supplementary data 5.

### Steps of running MR methods

#### Step 1: quality control of GWAS summary statistics

The aim of the quality control step is to identify a candidate set of SNPs with high quality for IV selection and MR analysis. In our analysis, we adopted several common QC measures for GWAS summary statistics, including

- Checking missingness. For each SNP, the required data information for performing MR analysis includes SNP identifier (we use rs number), effect allele, none effect allele, effect size, standard error, sample size (*N*), and *p*-value. SNPs missing any of the required information should be removed.
- Checking duplicates. Duplicated SNPs are SNPs with the same SNP identifier. We removed SNPs with duplicates to avoid any potential errors.
- Keeping unambiguous SNPs. We only involved unambiguous SNPs in our analysis, i.e., SNPs with the allele types A/G, A/C, T/G, or T/C.
- Removing poorly imputed SNPs. The imputed information score (Info) is a measure of the quality of the imputed SNPs. SNPs with Info *<* 0.9 are likely to be poorly imputed and were excluded from analysis. This QC step is applicable as long as the imputed information is available in GWAS summary statistics.
- Removing low minor allele frequency (MAF) SNPs. Low MAF SNPs are those with MAF below a certain threshold (e.g., 0.01 or 0.05). SNPs with low MAF were excluded from analysis as they are more prone to error. The QC threshold for MAF was chosen as 0.01 in our analyses. We will show later that the MR analysis results from different methods are not sensitive to the QC threshold for MAF. This QC step is applicable as long as MAF is available in GWAS summary statistics.
- Keeping SNPs in the set of HapMap 3 list. Because MAF or imputed information may be missing from the GWAS summary statistics, like LDSC, we restricted the analysis to a set of common and well-imputed SNPs in the HapMap 3 reference panel.
- Removing SNPs in the complex Major Histocompatibility Region (Chromosome 6, 26Mb *−* 34Mb).
- Removing SNPs with extremely large *χ*^2^. We removed SNPs with *χ*^2^ *> max*{80, N/1000} to reduce the undue influence of outliers on MR analysis results

After the QC step, GWAS datasets were formatted by retaining only the necessary data information for a set of SNPs that meet pre-determined quality control criteria. The retained data information typically includes the rs number, effect allele, non-effect allele, effect size, standard error, and *p*-value. It is important to note that we assume the phenotype and genotypes in GWASs are scaled to have a mean of zero and a variance of one. This scaling allows for the effect size and standard error to be calculated from *z*-scores and sample size, which can be more easily obtained from GWAS summary statistics.

The QC step is also important for methods like MR-APSS and CAUSE, which use SNPs across the genome to estimate nuisance parameters for their model.

#### Step 2: harmonizing SNP effects of the exposure and outcome

Performing MR analysis for an exposure-outcome trait pair requires harmonizing the effect estimates of each SNP to refer to the same allele. This is crucial for accurate MR analysis, as it ensures that the effect estimates for each SNP are comparable and can be combined to estimate the causal effect of the exposure on the outcome. To achieve this, we first checked the strands of the exposure and outcome alleles and flipped the outcome allele to the same strand as the exposure allele if they differ. We then checked the effect alleles of the exposure and outcome, and if they differed, we flipped the direction of the SNP-outcome effect to ensure that all effect estimates were aligned to the same allele. For example, if an SNP had an effect/non-effect allele of A/G in the exposure GWAS and C/T in the outcome GWAS, we first flipped the outcome allele to G/A. As the outcome GWAS presents the effect for the non-effect allele in the exposure GWAS, we then flipped the direction of the outcome effect to its opposite. Note that only unambiguous SNPs with allele types A/G, A/C, T/G, or T/C were considered, and any ambiguous SNPs were discarded from the analysis.

#### Step 3: IV selection and extract IV effects

After obtaining the harmonized summary dataset for each exposure-outcome trait pair, we began selecting instrumental variables (IVs) by identifying SNPs that were reliably associated with the exposure trait using a *p*-value threshold. To examine the robustness of MR methods to weak IV bias, we varied the *p*-value threshold for IV selection from 5 *×* 10*^−^*^8^ to 5 *×* 10*^−^*^5^. For those methods that require independence, we further used the Plink LD clumping procedure with a threshold of *r*^2^ = 0.001 and a window size of 1 Mb to obtain a set of nearly independent SNPs from the initial set of SNPs that passed the p-value threshold. It is important to note that we required each trait pair to be analyzed with a minimum of five IVs. The final dataset containing the summary data for the selected IV set was used as input for performing MR analysis, with the goal of estimating the causal effect of the exposure on the outcome.

While it is common practice to select independent IVs from the exposure dataset and obtain summary data from the outcome GWAS, we perform IV selection after harmonizing the exposure and outcome datasets. This approach may reduce IV loss due to LD clumping, as selected IVs may be absent from the outcome GWAS.

#### Step 4: run MR methods

The implementation details of the 15 compared methods are described as follows:

- IVW-fixed, IVW-random, Egger, Weighted-median, and Weighted-mode were performed using the legacy version of the TwoSampleMR R package with default options (https://github.com/MRCIEU/TwoSampleMR).
- RAPS wass performed using the mr.raps package without diagnostics by setting diagnostics=F.
- MRMix was performed using the MRMix package with defalut option (https://github.com/gqi/MRMix).
- MR-PRESSO was performed using the MRPRESSO package (https://github.com/rondolab/MR-PRESSO) using OUTLIERtest = TRUE, DISTORTIONtest = TRUE, SignifThreshold = 0.05, seed = 1234, and NbDistribution = 1000 options.
- MR-Robust was performed with *lmrob* in robustbase R package.
- MR-Lasso and MR-ConMix are performed using the *mr lasso* and *mr conmix* functions, respectively, with their default options in MendelianRandomization R package.
- cML-MA wad performed using R function of *mr cML* with default options in the MRcML R package. It is important to note that we did not compare the results of the Data perturbation (DP) versions of cML-MA in our analysis. This decision was based on the consideration that the default cML-MA version (without DP) is more time-efficient.
- MR-APSS was performed using the MR-APSS (https://github.com/YangLabHKUST/MR-APSS) R package.
- CAUSE was performed using the CAUSE (https://github.com/jean997/cause) R package.
- MR-CUE was performed using the MR.CUE (https://github.com/QingCheng0218/MR.CUE) R package.

### The choice of minor allele frequency threshold in quality control step

One of the QC measures for GWAS summary statistics was to exclude low MAF SNPs with the concern that they are more prone to error. Typically, studies use MAF thresholds of 0.01 or 0.05. To assess the impact of the MAF threshold and to determine an appropriate MAF threshold for MR, we conducted an analysis to explore the effect of the MAF threshold by varying the MAF threshold. Specifically, we considered the analysis of the six UK Biobank trait pairs used to evaluate causal effect estimation. We used MAF thresholds of 0.01 and 0.05 in the QC step for the summary statistics, and we then applied MR methods to the formatted summary datasets with different MAF QC thresholds. Results from different MR methods using different MAF thresholds are given in Sfig. S2. Our analysis shows that MR methods are generally not sensitive to the choice of MAF thresholds. However, to obtain more candidate IVs, we chose a threshold of 0.01 for MAF QC in our MR analysis.

### The choice of reference panel for LD clumping

All of the compared MR methods, except for MR-CUE, require independent or weakly correlated IVs. For those methods, an LD reference panel was used to perform LD clumping in the IV selection step. In contrast, MR-CUE allows for correlated IVs, and an LD reference panel is used to model the correlation between SNPs. To examine whether MR methods are sensitive to the choice of LD reference panel, we conducted a sensitivity analysis by comparing the results obtained using different reference panels. Specifically, we used an in-sample UK Biobank LD reference panel and the 1000 Genomes reference panel of European ancestry for the six UK Biobank trait pairs used to evaluate causal effect estimation. Both reference panels are of the same ancestry as the study population. We present the results from different MR methods using different LD reference panels in Supplementary Figure Sfig. S3. Our analysis shows that MR methods are generally not sensitive to the choice of LD reference panel as long as the panels are from the same ancestry.

## Data availability

The UK Biobank data are from UK Biobank resources under application number 30186. All GWAS summary statistics used in this study are downloadable at https://github.com/YangLabHKUST/MRbenchmarking. Supplementary Data 1-3, and 5 provide the references of these datasets.

## Code availability

The source codes to reproduce all the analyses can be accessed at the following location: https://github.com/YangLabHKUST/MRbenchmarking.

## Acknowledgements

We acknowledge the following grants: Hong Kong Research Grant Council grants nos. 16301419, 16308120, 16307221 and 16307322, Hong Kong University of Science and Technology Startup Grants R9405 and Z0428 from the Big Data Institute, Guangdong-Hong Kong-Macao Joint Laboratory grant no. 2020B1212030001 and the RGC Collaborative Research Fund grant no. C6021-19EF to C.Y., City University of Hong Kong Startup Grant 7200746 and Strategic Research Grant 21300423 to M.C.

