## supplemental notes and figures for "Benchmarking Mendelian Randomization methods for causal inference using genome-wide association study summary statistics"

#### Supplementary Figures S1-S21

#### Supplementary Tables S1-S2

**Table** S1: Summary of compared methods . . . . . . . . . . . . . . . . . . . . . . 2

### 1 Compared MR methods

In this study, we considered 15 summary-level data-based MR methods including IVW-fixed, IVW-random, Egger, RAPS, Weighted-median, Weighted-mode, MR-PRESSO, MRMix, cML-MA, MR-Robust, MR-Lasso, MR-CUE, CAUSE, MRAPSS and MR-ConMix. These methods all belong to the family of polygenic MR which uses variants from multiple genetic regions across the whole genome to estimate the causal effect of an exposure on an outcome. We categorized these methods into four groups: the inverse-variance weighted (IVW) class (IVW-fixed and IVW-random), outlier detection and removal methods (MR-PRESSO, MR-Lasso, and cML-MA), outlier-robust methods (Weighted-median, Weighted-mode, MR-Robust), and model-based methods (Egger, RAPS, MRMix, MR-ConMix, MR-CUE, CAUSE, and MR-APSS). To proceed with an overview of these methods, we first introduce the basic framework of MR and the assumptions that underlie it.

**Table** S1: Summary of compared methods

| **Method** | **Method category** | **IV validity assumption** | **R package** |
|---|---|---|---|
| IVW-fixed | IVW-class | All IVs are valid | *TwoSampleMR* |
| IVW-random | IVW-class | All IVs can be invalid affected by (mean zero) uncorrelated pleiotropy | *TwoSampleMR* |
| MR-Lasso | Outlier detection and removal | Plurality valid | *MendeianRandomization* |
| cML-MA | Outlier detection and removal | Plurality valid | *MRcML* |
| MR-PRESSO | Outlier detection and removal | Majority IVs can be invalid affected by (mean zero) uncorrelated pleiotropy | *MR-PRESSO* |
| Weighted-median | Outlier robust | Majority valid | *TwoSampleMR* |
| MR-Robust | Outlier robust | Plurality valid | *robustbase* |
| Weighted-mode | Outlier robust | Plurality valid | *TwoSampleMR* |
| Egger | Model based | All IVs can be invalid affected by (non-zero mean) uncorrelated pleiotropy | *TwoSampleMR* |
| RAPS | Model based | All IVs can be invalid affected by (mean zero) uncorrelated pleiotropy | mr.raps |
| MRMix | (Mixture) model based | Plurality valid | *MRMix* |
| MR-ConMix | (Mixture) model based | Plurality valid | *MendeianRandomization* |
| MR-APSS | (Mixture) model based | All IVs can be invalid due to pleiotropy or population stratification | *MRAPSS* |
| CAUSE | (Mixture) model based | Majority IVs not affected by correlated pleiotropy | *cause* |
| MR-CUE | (Mixture) model based | Majority IVs not affected by correlated pleiotropy | *MR.CUE* |

#### 1.1 The basic framework of MR

**Three IV assumptions** Mendelian randomization is a method that uses genetic variants as instruments to assess the causal effect of an exposure ($X$) on the outcome ($Y$) in the presence of unmeasured confounder $U$. To ensure the validity of an MR analysis, genetic variants used as IVs should be valid satisfying three IV assumptions. As illustrated in Figure S1, these assumptions can be summarized as follows:

- IV-I: The IV ($G_j$) is associated with the exposure ($X$) of interest;

- IV-II: The IV ($G_j$) is not associated with the confounders ($U$) of the exposure ($X$) and the outcome ($Y$);
- IV-III: The IV ($G_j$) affects the outcome ($Y$) exclusively through the exposure ($X$) of interest.

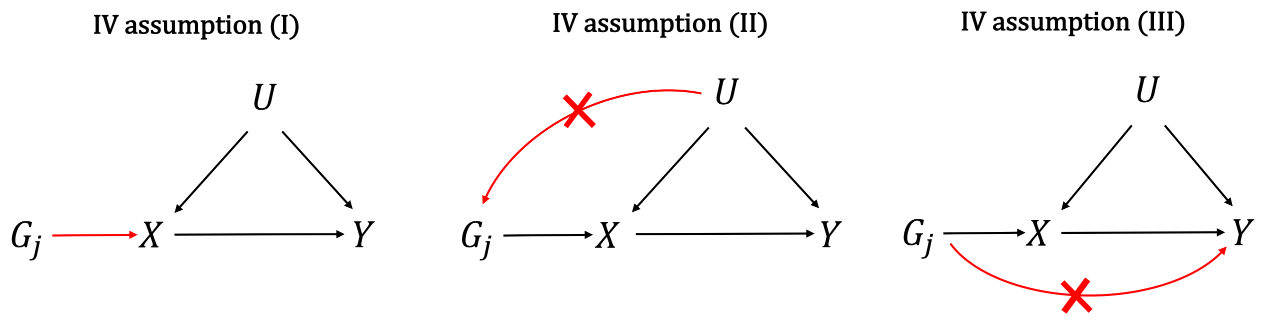

**Figure** S1: **The three IV assumptions**

**Assumed statistical model** Let $\{G_j\}_{j=1}^M$ be the $M$ SNPs used as IVs. We assume that all relationships between variables including $\{G_j\}_{j=1}^M, X, Y,$ and $U$ are linear and the $M$ SNPs used as IVs are independent of each other. We consider the following linear models for $X$ and $Y$,

$$X = \sum_{j=1}^{M} \gamma_j G_j + \eta_X U + \epsilon_{X,j}, \quad Y = X\beta + \sum_{j=1}^{M} \alpha_j G_j + \eta_Y U + \epsilon_{Y,j}, \tag{1}$$

where $\gamma_j$ is the effect of $G_j$ on $X$, $\alpha_j$ is the effect of $G_j$ on $Y$, $\beta$ be the causal effect of $X$ on $Y$ which is of interest, $\eta_X$ and $\eta_Y$ are the effect of confounder $U$ on $X$ and $Y$, respectively, and $\epsilon_{X,j}$ and $\epsilon_{Y,j}$ are the residual terms.

The linear model in Eq.1 can be represented as

$$X = \sum_{j=1}^{M} \gamma_j G_j + \eta_X U + \epsilon_{X,j}, \quad Y = \sum_{j=1}^{M} (\gamma_j \beta + \alpha_j) G_j + (\beta\eta_X + \eta_Y) U + \epsilon_{Y,j}, \tag{2}$$

If $G_j$ is a valid IV, we should have (IV-I) $\gamma_j \neq 0$, (IV-II) $G_j$ independent of $U$, and (IV-III) $\alpha_j = 0$. Therefore, we can obtain the following relationship for a valid IV,

$$\Gamma_j = \beta\gamma_j, \tag{3}$$

where $\gamma_j$ and $\Gamma_j$ represents the genetic effect of $G_j$ on $X$ and $Y$, respectively.

#### 1.2 Inverse variance weighted methods

Let us denote the GWAS estimates of the $j$-th SNP $G_j$ on the exposure and outcome as $\hat{\gamma}_j$ and $\hat{\Gamma}_j$, respectively, with standard errors $\hat{\sigma}_{X,j}$ and $\hat{\sigma}_{Y,j}$. Equation (3) indicates the causal effect can be estimated using the $j$-th SNP as the ratio of the SNP-outcome effect estimate to the SNP-exposure effect estimate:

$$\hat{\beta}_j = \frac{\hat{\Gamma}_j}{\hat{\gamma}_j}. \tag{4}$$

Assuming negligible estimation error of the SNP-exposure effect $\hat{\gamma}_j$ (No Measurement Error assumption or ***NOME assumption***), the standard error of the estimate can be obtained using the delta method:

$$\text{SE}(\hat{\beta}_j) = \sqrt{\frac{\hat{\sigma}^2_{Y,j}}{\hat{\gamma}^2_j}}.$$

**IVW-fixed** refers to the inverse variance weighted fixed-effects meta-analysis, which combines the ratio estimates from $M$ IVs to obtain a causal effect estimate under the framework of meta-analysis. However, in addition to the NOME assumption, two key assumptions are further needed: (1) all IVs are valid, and (2) there is no heterogeneity among the ratio estimates across IVs. Under these assumptions, the IVW estimator is therefore computed as a weighted mean of the ratio estimates of the $M$ IVs, as shown in Eq. (5):

$$\hat{\beta} = \frac{\sum_{j=1}^{M} \hat{\Gamma}_j \hat{\gamma}_j \hat{\sigma}^{-2}_{Y,j}}{\sum_{j=1}^{M} \hat{\gamma}^2_j \hat{\sigma}^{-2}_{Y,j}}. \tag{5}$$

The IVW estimator in Eq.(5) can be equivalently obtained as the slope of a weighted regression between the SNP-outcome effect estimates ($\hat{\Gamma}_j$) and the SNP-exposure effect estimates ($\hat{\gamma}_j$) with a fixed intercept of zero and inverse variance weights ($\hat{\sigma}^{-2}_{Y,j}$):

$$\hat{\Gamma}_j = \beta\hat{\gamma}_j + \epsilon_j, \quad \epsilon_j \sim \mathcal{N}(0, \hat{\sigma}^2_{Y,j}), \tag{6}$$

where $\epsilon_j$ is the error term. The variance of the IVW-fixed estimator is obtained as

$$\text{Var}(\hat{\beta}) = \frac{1}{\sum_j \hat{\gamma}^2_j \hat{\sigma}^{-2}_{Y,j}}. \tag{7}$$

**IVW-random** known as the inverse variance weighted fixed-effects meta-analysis, on the other hand, is a variation of the IVW-fixed estimator that incorporates residual heterogeneity into the model. This heterogeneity can arise due to various factors, such as unmeasured confounding factors like pleiotropy. To account for this heterogeneity, the IVW-random estimator incorporates a multiplicative factor ($\phi$) on the variance of the error term. As such, the regression equation in Eq. (6) becomes

$$\hat{\Gamma}_j = \beta\hat{\gamma}_j + \epsilon_j, \quad \epsilon_j \sim \mathcal{N}(0, \phi\hat{\sigma}^2_{Y,j}), \tag{8}$$

where $\phi$ is an overdispersion parameter and should be greater than 1. The inclusion of the overdispersion parameter will not affect the point estimate of the causal effect, meaning that the causal effect estimate of IVW-fixed and IVW-random will be the same as given in Eg. (5). However, the variance will be different, which is obtained as

$$\text{Var}(\hat{\beta}) = \frac{\hat{\phi}}{\sum_j \hat{\gamma}^2_j \hat{\sigma}^{-2}_{Y,j}}, \tag{9}$$

where $\hat{\phi}$ is the estimate of the dispersion parameter. In the presence of heterogeneity across IVs, $\hat{\phi}$ can be larger than 1 because of the overdispersion of genetic effects, making the standard error of IVW-random estimator larger than that of IVW-fixed.

#### 1.3 Outlier-robust methods

The second group of MR methods aims to obtain causal effect estimates that are robust to the presence of outlier invalid instrumental variables (IVs). These methods include the MR-Robust, Weighted-mode, and Weighted-median estimators.

**MR-Robust** modifies the inverse-variance weighting (IVW) regression model to provide robustness against outliers. Unlike the standard IVW methods which use the ordinary least squares for estimation, MR-Robust combines the robust regression technique MM-estimation with Tukey's bisquare loss function to obtain a more robust estimate of the causal effect. This approach allows us to put more weight on the more reliable ratio estimates and less weight on outlier estimates while still incorporating information from all included IVs.

**Weighted-median** aims to obtain the median of the ratio estimates of multiple IVs. Unlike the IVW estimator that computes a weighted mean of ratio estimates, which can be biased by the inclusion of invalid IVs, the weighted-median estimator is consistent even when up to 50% of the SNPs are invalid IVs. In other words, the weighted-median estimator only requires the ***majority*** (over 50%) of the SNPs to be valid IVs, which is referred to as the majority valid assumption. In our analysis, we used a more efficient weighted version of the median estimator, where the weights are equal to the inverse of the variance of the ratio estimates, rather than the simple median estimator, where each SNP is assigned equal weight.

**Weighted-mode** estimates the causal effect using the mode of the ratio estimates. In our analysis, we used a weighted version of the mode estimator, where the weights are equal to the inverse of the variance of the ratio estimates. The causal effect is estimated as the value that maximizes the normal kernel density of the ratio estimates. Compared to the weighted-median estimator, the weighted-mode estimator requires a weaker assumption of ***plurality validity*** to ensure that the set of valid IVs falls into the largest group.

Both the weighted-median and weighted-mode estimators are designed to be more robust in the presence of invalid IVs when compared to the IVW estimator. However, they may be less efficient than the IVW estimator.

#### 1.4 Outlier detection and removal methods

Outlier detection and removal methods treat invalid IVs as outliers and try to obtain reliable causal estimates by removing them from the analysis. Methods in this group include MR-Lasso, cML-MA, and MR-PRESSO.

**MR-Lasso** extends the IVW regression model by introducing an intercept ($\alpha_j$) to each of the IVs to account for pleiotropy and puts a penalty on the $L_1$-norm of the intercept terms. The model is formulated as follows:

$$\sum_j \hat{\sigma}_{Y,j}^{-2}(\hat{\Gamma}_j - \beta\hat{\gamma}_j - \alpha_j)^2 + \lambda \sum_j |\alpha_j|, \tag{10}$$

where $\alpha_j$ represents the direct pleiotropic effect of the $j$-th IV $G_j$ on the outcome that is not mediated by the exposure ($X$). With the lasso penalty applied to the intercept terms, their estimates tend to be shrunk toward zero. The SNPs with zero intercept estimates are considered as valid IVs. Then, the causal effect is estimated by fitting an IVW-fixed regression using the valid IVs. The sparsity level of the intercepts relies on a tuning parameter ($\lambda$) of the lasso penalty. In our analysis, we chose the optimal value of $\lambda$ by performing a search over a range of values that satisfy the heterogeneity stopping rule, as is implemented in the *mr_lasso* function of the 'MendelianRandomization' R package. However, we found that the implemented code for the heterogeneity stopping rule has a risk of stopping at some $\lambda$ values where no $\alpha_j$ is shrunk to zero, indicating the absence of valid IV. This gives null results for some exposure-outcome trait pairs.

Unlike MR-Lasso which uses the sparsity property of $L_1$ penalty for detection of invalid IVs, **cML-MA** considers the constrained maximum likelihood approach where $L_0$ penalty is introduced for identification of invalid IVs. Specifically, cML-MA tries to solve the following problem,

$$min \sum_j \hat{\sigma}_{X,j}^{-2}(\hat{\gamma}_j - \gamma_j)^2 + \hat{\sigma}_{Y,j}^{-2}(\hat{\Gamma}_j - \beta\gamma_j - \alpha_j)^2, \quad \text{subject to} \sum_j I(\alpha_j \neq 0) = K, \tag{11}$$

where $I(\cdot)$ is the indicator function and $K$ represents the unknown number of invalid IVs. In cML-MA, $K$ is selected through the Bayesian information criterion.

Both MR-Lasso and cML-MA rely on the ***plurality valid*** assumption that out of all groups of IVs having the same asymptotic ratio estimates of the causal effect, the largest group is the group of valid IVs. MR-Lasso further assumes no measurement error (NOME) on the SNP-exposure effect estimates.

**MR-PRESSO**, known as MR pleiotropy residual sum and outlier, is developed based on the framework of the IVW regression. It exploits the rationale that valid IVs will have small regression residuals and will be close to the regression line, while invalid IVs will significantly deviate from the regression line with a larger absolute value of residuals. The outlier test of MR-PRESSO is thus designed by comparing the observed residual sum of squares to the distribution of the expected residual sum of squares which are simulated under the null of no horizontal pleiotropy. To ensure validity, MR-PRESSO requires that at least 50% of the SNPs are valid IVs or have ***balanced pleiotropy*** in which case the average of a direct pleiotropic effect ($\alpha_j$) should be zero. Furthermore, it requires the ***InSIDE*** (Instrument Strength Independent of Direct Effect) assumption, meaning that $\gamma_j$ and $\alpha_j$ are independent of each other.

#### 1.5 Model based methods

The final group of methods aims to address the issue of invalid instrumental variables (IVs) in MR by using mixture component models or other modeling techniques. These methods include Egger, RAPs, MRMix, MR-ConMix, MR-CUE, CAUSE, and MR-APSS.

Egger and RAPS rely on the following model of invalid IVs:

$$\Gamma_j = \beta\gamma_j + \alpha_j, \tag{12}$$

where the genetic effect of the $j$-th IV on outcome ($\Gamma_j$) can be decomposed into a sum of indirect effect (causal effect of the exposure ($\beta\gamma_j$)) and a direct (pleiotropic) effect ($\alpha_j$). Here, $\alpha_j$ should be zero for a valid IV and deviate from zero for an invalid IV. The ***InSIDE*** assumption is satisfied if $\gamma_j$ and $\alpha_j$ are assumed to be independent of each other.

**Egger** regression is designed to address the issue of directional pleiotropy, which occurs when the average of the pleiotropic effects ($\alpha_j$) is non-zero. To achieve this, Egger modifies the

IVW regression by introducing a non-zero intercept ($\alpha_0$), as shown below:

$$\hat{\Gamma}_j = \beta\hat{\gamma}_j + \alpha_0 + \epsilon_j, \quad \epsilon_j \sim \mathcal{N}(0, \phi\hat{\sigma}^2_{Y,j}), \tag{13}$$

where $\epsilon_j$ is a normally distributed error term with mean zero and variance $\phi\hat{\sigma}^2_{Y,j}$, However, for the estimator to be consistent, the ***InSIDE*** assumption must be satisfied. Moreover, the ***NOME*** assumption is also required by Egger because it ignores the estimation error of the SNP-exposure effect estimates.

**RAPS** offers a distinct approach to Egger's method by addressing concerns that arise when performing MR, such as bias due to weak pleiotropic effects and large pleiotropic outliers. Specifically, RAPS uses a random effects model to account for the pleiotropic effects, allowing for variation of effects across different genetic variants under the ***InSIDE*** assumption. The pleiotropic effect ($\alpha_j$) is assumed to follow a normal distribution with a mean of zero and a variance of $\tau^2$,

$$\Gamma_j = \beta\gamma_j + \alpha_j, \quad \alpha_j \sim N(0, \tau^2). \tag{14}$$

For estimation, RAPS uses a profile likelihood approach to simultaneously estimate the causal effect $\beta$ and the variance term $\tau^2$, as given by the following equation,

$$\ell(\beta, \tau^2) = -\frac{1}{2}\sum_{j=1}^{p} \frac{(\hat{\Gamma}_j - \beta\hat{\gamma}_j)^2}{\hat{\sigma}^2_{X,j}\beta^2 + \hat{\sigma}^2_{Y,j} + \tau^2} + \log(\hat{\sigma}^2_{Y,j} + \tau^2). \tag{15}$$

Notably, this approach relaxes the NOME assumption since the likelihood function accounts for the estimation error of $\hat{\gamma}_j$. To further reduce the influence of large pleiotropic outliers, RAPS employs a robust loss function, such as Tukey's weight loss function or Huber's loss function, which down weights the influence of outliers in the data and helps mitigate their impact on the estimated causal effect

Other methods belonging to this group, such as MRMix, MR-ConMix, MR-CUE, CAUSE, and MR-APSS, are all MR approaches that employ different mixture component models to address the potential issue of invalid instruments. The basic idea behind these methods is to assume that the genetic instruments are a mixture of two or more components, where one component contains causal signal used for causal inference and one or more components are used to account for IV invalidity. However, the specific models and algorithms used by each method are different.

**MRMix** employs a four-component mixture model to model the potential effects of IVs on the exposure and outcome ($\gamma_j$, $\Gamma_j$). The four components of the mixture model correspond to four different groups of IVs: valid IVs, invalid IVs violating the IV assumption (III) by preserving a pleiotropic effect on the outcome, and other two types of extremely weak IVs that violate IV assumption (I) by having no effect on the exposure:

- group 1: $\Gamma_j = \beta\gamma_j$, for $G_j$ is a valid IV satisfying the three IV assumptions;
- group 2: $\Gamma_j = \gamma_j\beta + \alpha_j$, $\begin{bmatrix}\gamma_j \\ \alpha_j\end{bmatrix} \sim \mathcal{N}\left(\begin{bmatrix}0 \\ 0\end{bmatrix}, \begin{bmatrix}\sigma^2 & \theta \\ \theta & \tau^2\end{bmatrix}\right)$, for $G_j$ is an invalid IV violate the IV assumption (III);
- group 3: $\gamma_j = 0$ and $\Gamma_j \sim \mathcal{N}(0, \tau^2)$, for $G_j$ is extremely weak IV and have a direct effect on the outcome;

- group 4: $\gamma_j = 0$ and $\Gamma_j = 0$, for $G_j$ is extremely weak IV and have no effect on outcome.

Notably, for the set of invalid IVs with pleiotropic effect, MRMix allows $\gamma_j$ and $\alpha_j$ to be correlated violating the InSIDE assumption; MRMix also allows a set of extremely weak IVs which are not associated with the exposure. While MRMix is flexible in its ability to model IV invalidity due to pleiotropy or weak IVs, it requires a large number of instruments to precisely identify the group of valid instruments and improve the accuracy of causal effect estimation. Moreover, to distinguish between the component of valid IVs and that of invalid IVs, MRMix further requires the ***plurality valid*** assumption. This assumption is necessary to identify the group of valid instruments and estimate the causal effect accurately.

**MR-ConMix** takes a simpler model compared to MRMix by dividing IVs into two groups: valid IVs and invalid IVs. Specifically, if $G_j$ is valid, its ratio estimate $\hat{\beta}_j$ is assumed to be normally distributed with mean $\beta$ (the true causal effect) and variance $se(\hat{\beta}_j)^2$, and if $G_j$ is invalid, $\hat{\beta}_j$ is assumed to be normally distributed with mean $\beta_j$ and variance $se(\hat{\beta}_j)^2$, and $\beta_j$ is further assumed to have a distribution of $\mathcal{N}(0, \psi^2)$:

- $\hat{\beta}_j \sim \mathcal{N}(\beta, se(\hat{\beta}_j)^2)$, if $G_j$ is valid;
- $\hat{\beta}_j \sim \mathcal{N}(\beta_j, se(\hat{\beta}_j)^2)$ with $\beta_j \sim \mathcal{N}(0, \psi^2)$, if $G_j$ is invalid.

Here the parameter $\psi^2$ accounts for the uncertainty of the asymptomatic value of the ratio estimates ($\beta_j$) of invalid IVs. Under these assumptions, a profile likelihood approach is then used for the estimation of the causal effect. Similar to MRMix, MR-ConMix also requires the ***plurality valid*** assumption to ensure consistency of the causal effect estimate.

Two practical issues should be noted when using this method. The first issue is that $\psi$ is a user-specified parameter. The optimal choice of $\psi$ may vary depending on the specific research question, and a sensitivity analysis may be preferred in practice. However, due to the analysis burden, we chose to use the default value of $\psi$ for all the trait pairs tested, which is set as 1.5 times the standard deviation of the ratio estimates. While using default values can simplify the analysis process, it is important to note that the optimal value of $\psi$ may vary depending on the specific research question, and using a single default value may not be appropriate in all cases. The second issue is that the confidence interval given by MR-ConMix is not guaranteed to be symmetric or a single range of values.

**CAUSE** is proposed to address invalid IVs due to correlated pleiotropy which occurs when IVs affect both the exposure and outcome through a shared heritable factor. Specifically, CAUSE divides IVs into two groups: a group of IVs affected by correlated pleiotropy, and a group of IVs not affected by correlated pleiotropy which will be used for causal inference. The two groups of IVs are modeled as follows,

- group 1: $\Gamma_j = \beta\gamma_j + \eta\gamma_j + \alpha_j$, for $G_j$ is affected by correlated pleiotropy.
- group 2: $\Gamma_j = \beta\gamma_j + \alpha_j$, for $G_j$ is not affected by correlated pleiotropy;

where $\beta\gamma_j$ is the causal effect of $G_j$ on the outcome through the exposure, $\alpha_j \perp\!\!\!\perp \gamma_j$ represents the effect of uncorrelated pleiotropy, $\eta$ is a scalar, and $\eta\gamma_j$ represents the IV to outcome effect that is mediated by a shared confounding factor, termed as correlated pleiotropy.

Under the CAUSE model, all IVs are allowed to be invalid preserving an uncorrelated pleiotropic effect ($\alpha_j$). However, only a small proportion of SNPs (less than 50%) could be affected by correlated pleiotropy. This is essential to ensure the model's identifiability. By accounting for both uncorrelated and correlated pleiotropy, the CAUSE method tries to mitigate the risk of false positives in MR and enhance the accuracy of causal inference.

For estimation, CAUSE adopts a two-step procedure. In the first step, the distribution of IV to exposure effect ($\gamma_j$) and the uncorrelated pleiotropic effect ($\alpha_j$) are estimated using genome-wide summary statistics where they are assumed to follow a mixture of bivariate normal distribution. In the second step, CAUSE conducts a model comparison between a shared model ($\beta$ is fixed at zero) and a causal model ($\beta$ allows to be nonzero). The shared model assumes that there is no causal effect of the IV on the outcome through the exposure, while the causal model allows for a nonzero causal effect. The model comparison is achieved by comparing the expected log pointwise posterior density (ELPD) of the two models. The model with the higher ELPD is selected as the better model with posteriors that predict the data better.

**MR-CUE** (MR with Correlated horizontal pleiotropy Unraveling shared Etiology and confounding) is a method similar to CAUSE that accounts for correlated pleiotropy in MR. Like CAUSE, MR-CUE also divides IVs into two groups based on the presence or absence of correlated pleiotropy. However, MR-CUE differs from CAUSE in several key aspects.

First MR-CUE and CAUSE have different model specifications on correlated pleiotropy. CAUSE models the effect of correlated pleiotropy as $\eta\gamma_j$, i.e., the IV to outcome effect mediated by shared confounder (Figure S?-a). This is based on the assumption of a common confounder between the exposure and outcome. In contrast, MR-CUE aims to account for more complex situations with multiple confounders by modeling the effect of a confounder set on the outcome as a sum of two components, $\delta\gamma_j + \theta_j$ ($\delta$ is a scalar) (Figure S?-b). The first component summarizes the IV-shared confounding effect, and the second component represents the IV-specific perturbation effects of confounders. The assumed mixture model of MR-CUE is given as follows,

- $\Gamma_j = \beta\gamma_j + \alpha_j$, if $G_j$ is not affected by correlated pleiotropy;
- $\Gamma_j = \beta\gamma_j + (\delta\gamma_j + \theta_j) + \alpha_j$, if $G_j$ is affected by correlated pleiotropy.

where the pleiotropic effect terms $\theta_j$ and $\alpha_j$ are assumed to independent of the IV-exposure effect $\gamma_j$ and are draw from a normal distribution with zero mean.

Additionally, MR-CUE allows for correlation between the IVs, while CAUSE and other methods treat the IVs as independent. As a result, MR-CUE does not require a genome reference panel to perform linkage disequilibrium (LD) clumping in data preprocessing. Instead, to model the correlation between IVs, MR-CUE requires an LD reference panel, which is not required by other methods.

Finally, MR-CUE estimates the parameters with a Bayesian hierarchical model and performs inference via Gibbs sampling, while CAUSE estimates posterior distributions via adaptive grid approximation and performs inference by model comparison.

**MR-APSS** is a unified approach to MR accounting for IV invalidity due to pleiotropy and population stratification etc. The method uses a foreground-background model that separates observed SNP effect sizes into background and foreground components. The background component accounts for confounding factors that could induce a genetic correlation between the exposure and outcome, as well as factors that could lead to a correlation between estimation errors. In contrast, the foreground component is used for causal inference while accounting for uncorrelated pleiotropy. The assumed model for MR-APSS is given as follows:

$$\begin{pmatrix}\hat{\gamma}_j\\ \hat{\Gamma}_j\end{pmatrix} = Z_j \underbrace{\begin{pmatrix}\gamma_j\\ \beta\gamma_j + \alpha_j\end{pmatrix}}_{\text{Foreground}} + \underbrace{\begin{pmatrix}u_j\\ v_j\end{pmatrix} + \begin{pmatrix}e_j\\ \xi_j\end{pmatrix}}_{\text{Background}}, \quad j = 1, \ldots, M. \tag{16}$$

where $u_j$ and $v_j$ are the polygenic effects of SNP $j$ on $X$ and $Y$, $\epsilon_j$ and $\xi_j$ are the estimation errors of SNP effect sizes, The sum of $u_j$ and $\epsilon_j$ corresponds to the background effect on $X$ and the sum of $v_j$ and $\xi_j$ corresponds to the background effect on $Y$, $\gamma_j$ is the remaining SNP effect on exposure $X$ as the instrument strength, $\alpha_j$ is the direct SNP effect on outcome $Y$, $Z_j$ is a indicator that takes the value of 1 if $G_j$ carries foreground effect and 0 otherwise. In brief, MR-APSS assumes all IVs can be invalid having background effects but only a subset of them carrying foreground effects, and the set of IVs carrying foreground effects will be used for causal inference.

The MR-APSS model assumes that all instruments can be invalid, carrying background effects, but only a subset of them carries foreground effects that can be used for causal inference. The background model parameters are estimated using the LDSC assumption, which can be pre-estimated using genome-wide summary statistics. The foreground model relies on the InSIDE assumption, which ensures that IV strength and the direct effect on the outcome are independent of each other. The causal effect and other foreground model parameters are estimated using a variational EM algorithm. Importantly, MR-APSS is the only method among these compared methods that correct for selection bias in the two-sample setting. This feature allows for more accurate and reliable causal effect estimation in MR studies.

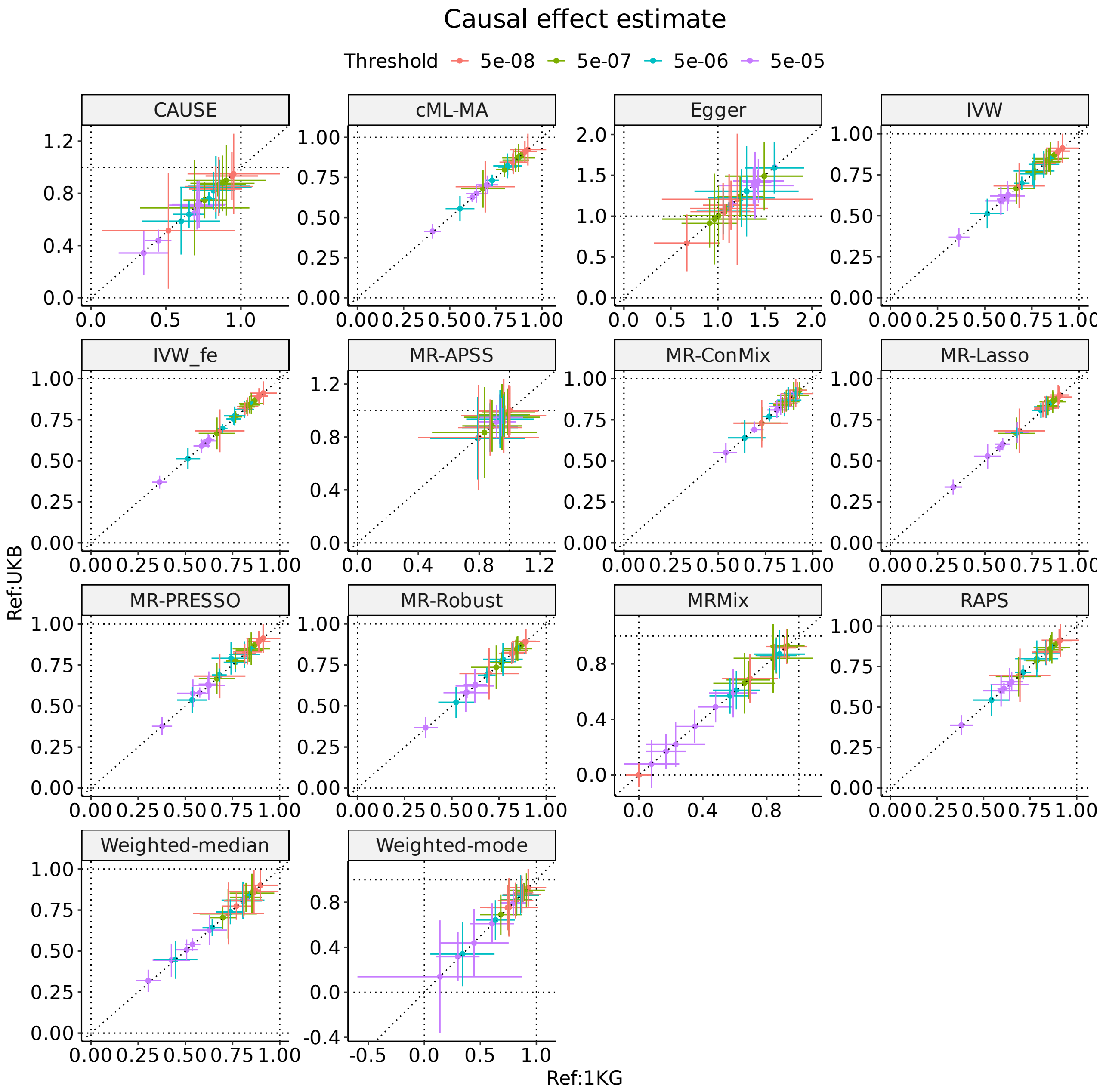

**Figure** S2: **Results from different MR methods using different MAF (Minor allele frequency) thresholds for data-preprocessing.** The X-axis represents estimates using MAF threshold of 0.01, and The X-axis represents estimates using MAF threshold of 0.05.

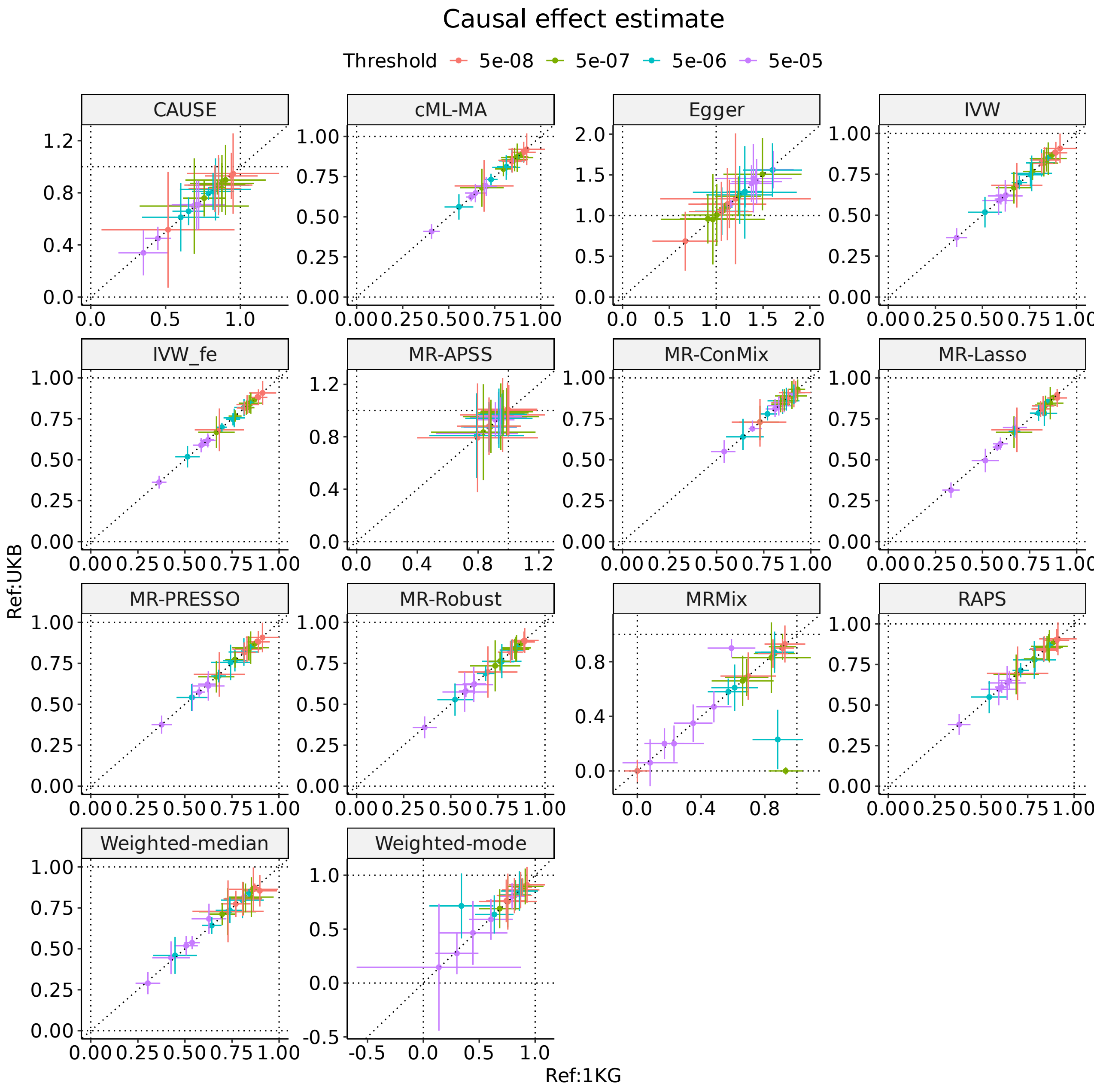

**Figure** S3: **Results from different MR methods using different LD reference panel for LD clumping**. The X-axis represents estimates using 1000 genome samples of European ancestry as a reference panel, and The X-axis represents estimates using 1000 samples of UK biobank as a reference panel.

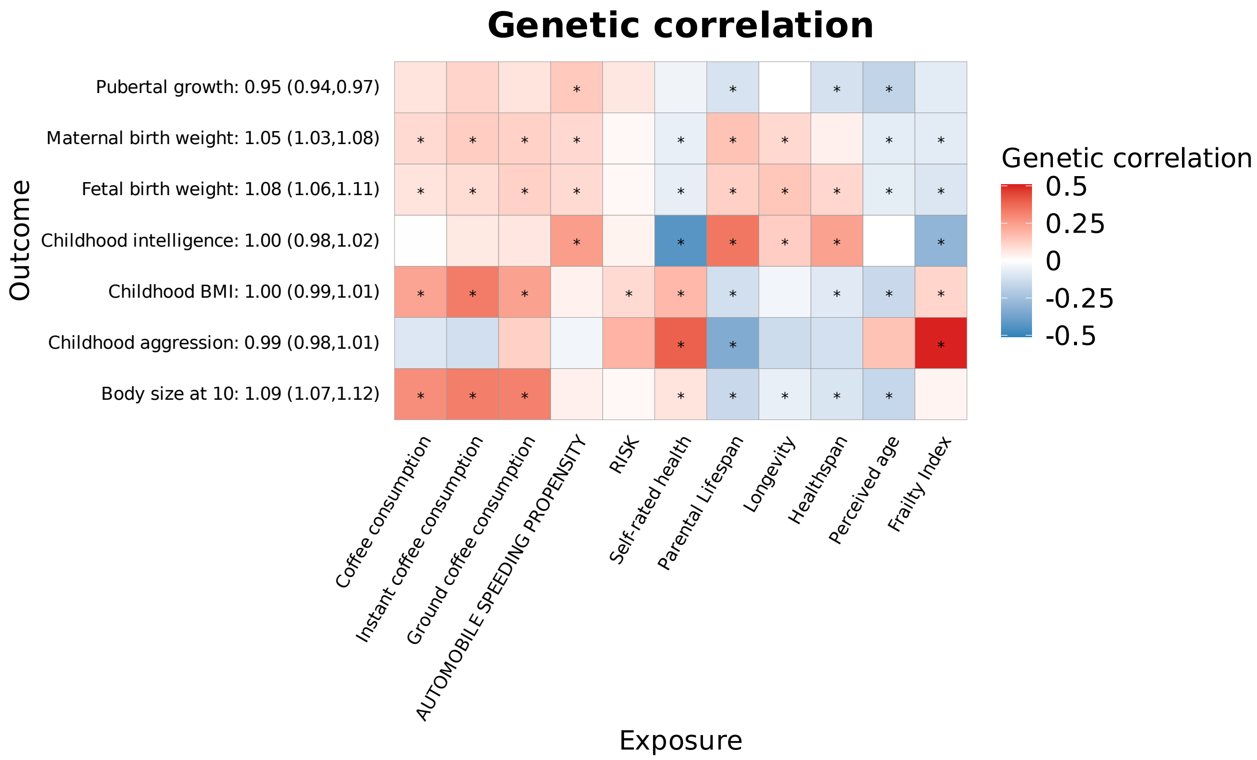

**Figure S4: LDSC results for the 77 negative control trait pairs**
